# Impact of COVID-19 pandemic and anti-pandemic measures on tuberculosis, viral hepatitis, HIV/AIDS and malaria – a systematic review

**DOI:** 10.1101/2022.02.10.22270782

**Authors:** Barbora Kessel, Torben Heinsohn, Jördis J Ott, Jutta Wolff, Max J Hassenstein, Berit Lange

## Abstract

COVID-19 pandemic puts an enormous strain on health care systems worldwide and may have a detrimental effect on prevention, treatment and outcomes of tuberculosis (TB), viral hepatitis, HIV/AIDS and malaria, whose ending is part of the United Nations 2030 Agenda for Sustainable Development. We conducted a systematic review of scientific and grey literature in order to collect wide-ranging evidence with emphasis on quantification of the projected and actual indirect impacts of COVID-19 on the four infectious diseases with a global focus. We followed PRISMA guidelines and the protocol registered for malaria (CRD42021234974). We searched PubMed, Scopus, preView (last search: January 13, 2021) and websites of main (medical) societies and leading NGOs related to each of the four considered infectious diseases. The identified modelling studies warned about under-diagnosis (TB), anti-retroviral therapy interruption/decrease in viral load suppression (HIV), disruptions of insecticide-treated nets (ITN) distribution and access to effective treatment (malaria), and treatment delays and vaccination interruptions (viral hepatitis). The reported disruptions were very heterogeneous both between and within countries. If observed at several points in time, the initial drops (partly dramatic, e.g. TB notifications/cases, or HIV testing volumes decreased up to -80%) were followed by a gradual recovery. However, the often-missing assessment of the changes against the usual pre-pandemic fluctuations hampered the interpretation of less severe ones. Given the recurring waves of the pandemic and the unknown mid- to long-term effects of adaptation and normalisation, the real consequences for the fight against leading infectious diseases will only manifest over the coming years.

## 1. Introduction

COVID-19 pandemic poses a direct threat to healthy individuals. Moreover, the efforts dedicated to its resolution along with the pervasive measures taken have a collateral impact on the health and care of persons with other diseases. On the one hand, there are clinical interrelations with either a higher infection risk or severity of COVID-19 in individuals living with another disease. On the other hand, the capacity of health care systems is stretched thin by the new challenges and resource adaptations needed to combat COVID-19, e.g. [1-3]. Interruptions in care can aggravate existing chronic conditions, or delay an early detection of new cases, which in turn can lead to worse treatment outcomes in the long term and/or to an increased risk of further transmission of the disease. As of 2019, tuberculosis (TB), malaria, HIV/AIDS and viral hepatitis (counting also cirrhosis and liver cancer secondary to hepatitis) were among the 20 leading causes of years of life lost globally [4]. Their ending is part of goal 3.3 of the United Nations 2030 Agenda for Sustainable Development [5]. Specific strategies exemplifying this goal in terms of reductions in new cases and deaths have been developed [6-9]. All these strategies stress the need for early detection, an increase in diagnosis and treatment coverage, and in vaccination coverage for TB and hepatitis. Sustainable funding, good management of supply chains, surveillance, and reaching populations with poor access to health services are crucial in the process [7, 10]. The progress made towards reaching the 2030 Sustainable Development Goals is now threatened in the wake of the COVID-19 pandemic’s global strain on resources. In our systematic review, we collect all evidence published in scientific and grey literature that describes the impact of COVID-19 and the pandemic control measures on prevention, treatment and outcomes of TB, viral hepatitis, HIV/AIDS and malaria. Our focus is on quantitatively assessed impact, such as changed numbers of case notifications, screening tests, vaccinations, or quantitatively described changes in service availability. Since the potential effect of the disruptions in care on new cases and mortality occur mostly with a delay (e.g. [11]), we also summarise projected effects on these two endpoints as reported from modelling studies. Previous systematic and non-systematic reviews focused on only HIV care [12], or developments in a specific region [13, 14], or are limited to scoping evidence without providing actual ranges of the quantitative developments [15]. We further the existing knowledge by synthesizing wide-ranging evidence with emphasis on quantification of the projected and actual indirect impacts of COVID-19 on TB, viral hepatitis, HIV and malaria with a global focus. The collected published evidence covers mostly the first six months of the pandemic.

## 2. Methods

The reporting of this review follows the PRISMA 2020 checklist [16] (see S2) and the study protocol registered for malaria (CRD42021234974). For transparency, S1 section 3 lists post-hoc amendments to the protocol.

### 2.1 SSearch strategy and selection criteria

We searched PubMed, Scopus and preView for COVID-19 or SARS-CoV-2 and at least one of hepatitis, tuberculosis, HIV, or malaria (last search date: January 13, 2021), without language restrictions. After de-duplication, we sorted the records into five groups: HIV, tuberculosis, malaria, viral hepatitis, and records not assigned to any of the diseases (see S1 section 1). Preprints with available full-texts and all but the latest versions of a preprint counted as a duplicate. We also conducted a grey literature search by manually searching websites of main (medical) societies and leading NGOs related to each of the four considered infectious diseases (see S1 section 2). Further, we screened references of included publications for additional relevant sources.

A study (a preprint, a published scientific paper, a report of the professional societies or program providers/funders, a conference abstract) was eligible for inclusion if it reported on the effect of COVID-19 outbreak on the treatment/prevention programme for at least one of TB, HIV, viral hepatitis, or malaria. In particular,

- on (rates of) newly diagnosed cases, or on number of deaths,
- on the accessibility of the services and access to medication from the perspective of individuals in, or eligible for, the programme,
- on availability of the services as provided by the providers,
- on changes in adherence to treatment and on clinical outcomes of services.

We also included modelling studies reporting the potential impact of the above effects on incidence or deaths for at least one of the considered diseases. We excluded studies that did not report the period to which the presented data referred, or did not state the source of the data. Similarly, the pre-pandemic comparison period had to be described, except for surveys, where we accepted questions using an implicit comparison (e.g. with the wording “due to COVID-19”, or “under current travel restrictions”).

Screening was done by two authors independently (TB: TH and BL, hepatitis: JJO and BK, HIV/AIDS: BK and MH, malaria: JW and TH) and inclusion of records was based on mutual agreement, or if unreachable, decided by a third author (BL). Studies in languages other than English were translated using Google Translate (https://translate.google.com/).

### 2.2 Data analysis

The risk of bias was assessed by two authors (TB: TH, BL, HIV: BK, MH, malaria: JW, TH, hepatitis: JJO, BK), using three different tools (see S1 section 5) according to the type of the study (modelling study, survey, other quantitative studies) based on [17-19]. We present the results graphically using the *robvis* package [20] in R. We did not exclude any study due to a high risk of bias, but we describe the resulting limitations in the discussion and we make a distinction between different data sources (official health records, surveys, grey literature) when summarising the results. From each included publication, we extracted results from the main analysis related to all relevant endpoints, together with the description of the studied population, country/region, study period and the used pre-pandemic comparison intervals. We extracted proportions (incl. the denominator if available), absolute numbers, or odds-ratios with the reported 95% confidence intervals (see S1 sections 7-10). If necessary, we used an online plot digitizer [21] to extract approximate numbers from published plots. The data extraction was done by one author and checked by another (TB: TH, BL, HIV: MH, BK, malaria: JW, TH, hepatitis: JJO, BK). We group the findings by endpoints and further by regions. In order to maximize the number of comparable results for each endpoint, we used the explicitly reported absolute numbers to calculate certain rates or percent changes as reported in studies providing less detailed information. We used R, Version 4.1.1 [22] for any necessary calculations. S1 Tables 7-22 also show the considered effect measures for each endpoint. For graphical presentation, we use simple line graphs (package *ggplot2* [23] in R) and forest plots (package *forestplot* [24] in R). For the latter, we enhance the extracted proportions with 95% Agresti-Coull confidence intervals (library *binom* [25] in R). Due to the heterogeneity of the results, we refrain from conducting a meta-analysis. We discuss the heterogeneity sources in section 4.

## 3. Results

**Fig. 1** shows the number of studies assessed, screened in full-text and finally included in our review. Further details about the search results and “near misses” are in S1 section 4. We found seven modelling studies for TB [26-32], ten for HIV [31, 33-41], three for hepatitis [42-44] and seven for malaria [31, 41, 45-49]. The main risk-of-bias concern was the missing uncertainty analysis, which was the case for several modelling studies. Surveys were sparse for malaria (three [50-52]) and relatively abundant for HIV (twenty-nine [51, 53-80]), with the numbers for hepatitis [68, 75, 81-84] and TB [51, 62, 85-88] in between. The majority of surveys did not report on missing data within the questionnaires. Studies related to TB, viral hepatitis and malaria often did not disclose the questionnaires, nor did they report on their testing. In contrast, for HIV, the dominant reason for concern was the representativeness of the sample/population, since the majority of the studies relied on convenience samples and respondents hired through social media. Regarding studies and reports based on patient, hospital or other official records (TB: [26, 28, 29, 89-111], viral hepatitis: [112-123], malaria: [41, 52, 124-132], HIV: [110, 133-163]), several single-centre studies had a high risk of bias, since our assessment was done from the global perspective. In a few other cases, reporting only a single, selected aggregated number gave rise to concerns. We did not assess the risk of bias for a press article [123] relevant for viral hepatitis. The details of the risk-of-bias assessment are shown in S1 section 6.

**Fig. 1.**
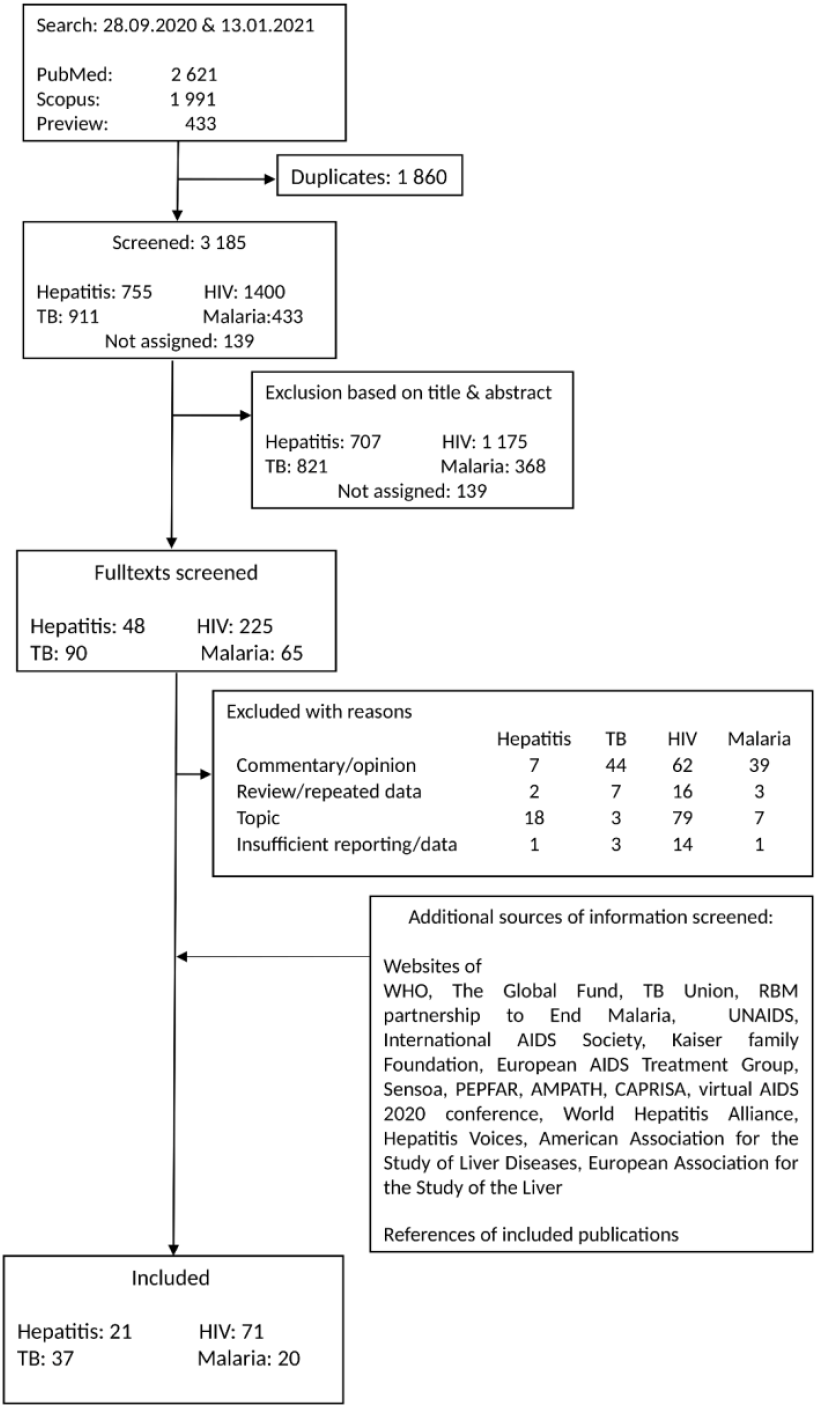
Flow chart. The final numbers of included records are sums of the records identified through the systematic search and the grey literature search. For details on the grey literature search see S1 section 2. Further details on the search results and “near misses” are in S1 section 4. Note that for malaria, two publications [130, 164] have the same contents despite different identifiers, and we decided to refer only to [130] throughout the text. Further, two studies on HIV from Japan[139, 148] evaluated the same data, so we will drop reporting on [148].

Due to the high variety of reported outcomes (**Fig. 2**), we did not assess the publication bias formally. For different indicators, we found reports of both positive and negative impact. No impact was reported as well. Nevertheless, we assume that larger negative impacts were considered more interesting and worth quick publication. **Fig. 2** gives an overview of all relevant outcomes identified in the literature.

**Fig. 2.**
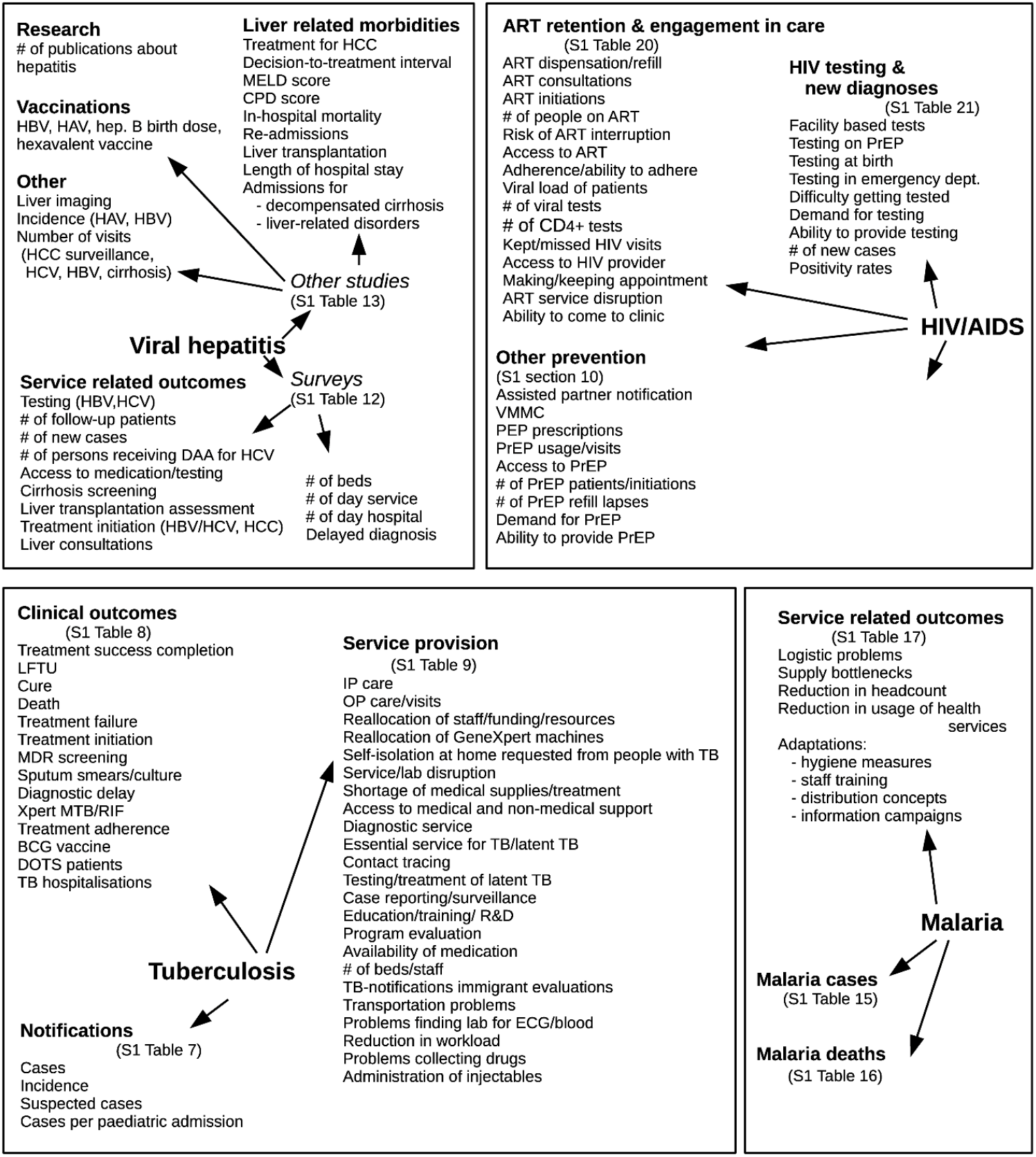
Overview of relevant outcomes identified in the literature. The detailed effects can be found in supplementary material as indicated

Besides examining a wide range of disruption scenarios, the modelling studies often identified single aspects of prevention and treatment programmes, disruption of which leads to the largest effects on new cases and deaths **(Table 1**). The observed disruptions of these most influential aspects are summarized in **Table 2** (TB, HIV, viral hepatitis) and **Table 3** (malaria). In the following subsections, we describe the main findings for each of the four infectious diseases separately. The detailed results are collected in S1 section 7 (TB), section 8 (viral hepatitis), section 9 (malaria) and section 10 (HIV). Note that if we refer loosely to the year 2020, we refer mostly to its first semester, since most studies covered March-June 2020, and almost none reported beyond August 2020.

**Table 1.**
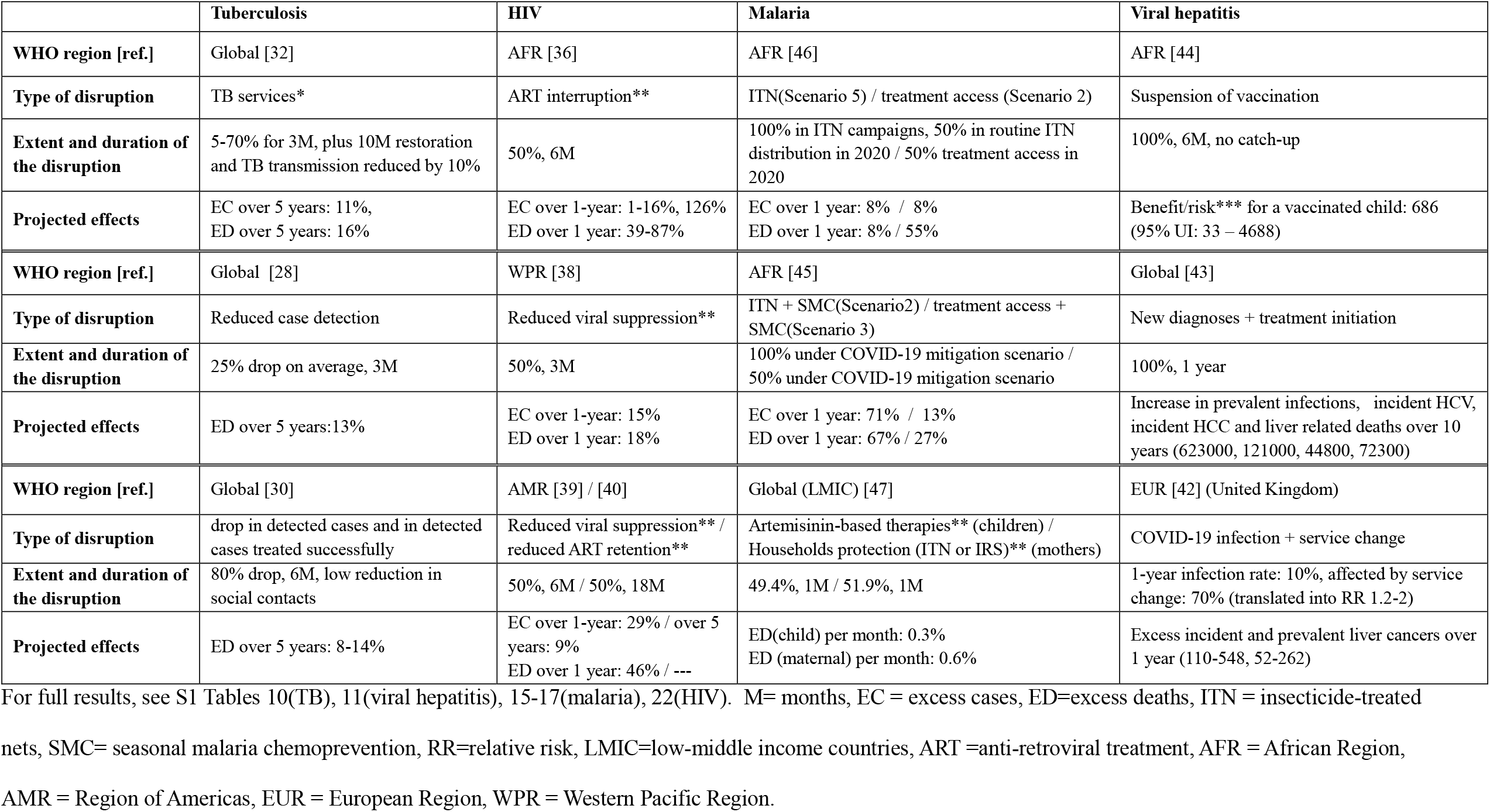
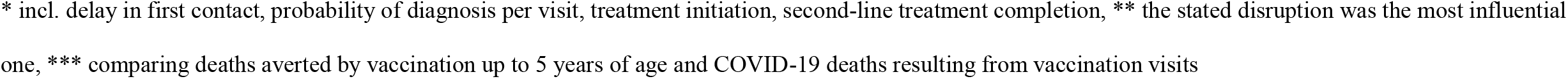
Overview of selected modelling results (three per each infectious disease) regarding indirect impact of COVID-19 on tuberculosis, malaria, HIV/AIDS and viral hepatitis.

|  | **Tuberculosis** | **HIV** | **Malaria** | **Viral hepatitis** |
| --- | --- | --- | --- | --- |
| **WHO region [ref.]** | Global [32] | AFR [36] | AFR [46] | AFR [44] |
| **Type of disruption** | TB services* | ART interruption** | ITN(Scenario 5) / treatment access (Scenario 2) | Suspension of vaccination |
| **Extent and duration of the disruption** | 5-70% for 3M, plus 10M restoration and TB transmission reduced by 10% | 50%, 6M | 100% in ITN campaigns, 50% in routine ITN distribution in 2020 / 50% treatment access in 2020 | 100%, 6M, no catch-up |
| **Projected effects** | EC over 5 years: 11%,<br>ED over 5 years: 16% | EC over 1-year: 1-16%, 126%<br>ED over 1 year: 39-87% | EC over 1 year: 8% / 8%<br>ED over 1 year: 8% / 55% | Benefit/risk*** for a vaccinated child: 686<br>(95% UI: 33 – 4688) |
| **WHO region [ref.]** | Global [28] | WPR [38] | AFR [45] | Global [43] |
| **Type of disruption** | Reduced case detection | Reduced viral suppression** | ITN + SMC(Scenario2) / treatment access + SMC(Scenario 3) | New diagnoses + treatment initiation |
| **Extent and duration of the disruption** | 25% drop on average, 3M | 50%, 3M | 100% under COVID-19 mitigation scenario / 50% under COVID-19 mitigation scenario | 100%, 1 year |
| **Projected effects** | ED over 5 years:13% | EC over 1-year: 15%<br>ED over 1 year: 18% | EC over 1 year: 71% / 13%<br>ED over 1 year: 67% / 27% | Increase in prevalent infections, incident HCV, incident HCC and liver related deaths over 10 years (623000, 121000, 44800, 72300) |
| **WHO region [ref.]** | Global [30] | AMR [39] / [40] | Global (LMIC) [47] | EUR [42] (United Kingdom) |
| **Type of disruption** | drop in detected cases and in detected cases treated successfully | Reduced viral suppression** / reduced ART retention** | Artemisinin-based therapies** (children) / Households protection (ITN or IRS)** (mothers) | COVID-19 infection + service change |
| **Extent and duration of the disruption** | 80% drop, 6M, low reduction in social contacts | 50%, 6M / 50%, 18M | 49.4%, 1M / 51.9%, 1M | 1-year infection rate: 10%, affected by service change: 70% (translated into RR 1.2-2) |
| **Projected effects** | ED over 5 years: 8-14% | EC over 1-year: 29% / over 5 years: 9%<br>ED over 1 year: 46% / --- | ED(child) per month: 0.3%<br>ED (maternal) per month: 0.6% | Excess incident and prevalent liver cancers over 1 year (110-548, 52-262) |

For full results, see S1 Tables 10(TB), 11(viral hepatitis), 15-17(malaria), 22(HIV). M= months, EC = excess cases, ED=excess deaths, ITN = insecticide-treated nets, SMC= seasonal malaria chemoprevention, RR=relative risk, LMIC=low-middle income countries, ART =anti-retroviral treatment, AFR = African Region, AMR = Region of Americas, EUR = European Region, WPR = Western Pacific Region.
\* incl. delay in first contact, probability of diagnosis per visit, treatment initiation, second-line treatment completion, \*\* the stated disruption was the most influential one, \*\*\* comparing deaths averted by vaccination up to 5 years of age and COVID-19 deaths resulting from vaccination visits

**Table 2.** Brief overview of observed impacts of COVID-19on selected endpoints for TB, HIV and viral hepatitis.

|  | WHO region |  |  |  |  |
| --- | --- | --- | --- | --- | --- |
|  | AFR | AMR | EUR | SEAR | WPR |
| **Tuberculosis** |  |  |  |  |  |
| Notifications/cases<sup>a</sup> | -78.6% to -12.6% [29, 94, 99, 109, 110, 164] | -80.8% to -2.6%[90, 92, 109] | -60% to +6.3%[108, 109] | -78% to -9.8%[26, 28, 91, 98, 102] | -48.6% to -5% [28, 87, 93, 95, 97, 100, 101, 103-106, 109] |
| Treatment success/completion rates<sup>b</sup> | -16.8% to 0.2%[110] |  | +1%[108] |  | -10% to -2%[95, 106] |
| Diagnostic delay[days]<br>Median(IQR) |  |  |  |  | Pandemic: 18.5(9-33), pre-pandemic: 15 (6-29)[97] |
| **HIV** |  |  |  |  |  |
| ART<br>• ART refill problems<br>• adherence to ART during pandemic<br>• ART dispensation | • 3%[57]<br>• Decreased ability to adhere: 14%[65]<br>• Coverage: 97%[150] | • 6%,7%, 17%[53, 72, 80]<br>• Decreased ability to adhere: 5%[72], adherence lower [53], better [61] (beginning of lockdown)<br>• --- | • ---<br>• ---<br>• -23.1%[153] | • ---<br>• ---<br>• approx. 50% reduction (weekly)[163] | • 18%[77]<br>• ---<br>• --- |
| Viral load suppression in lockdown |  | OR 1.31 for unsuppressed[157], sustained[141] | Sustained [140] |  |  |
| **Viral hepatitis** |  |  |  |  |  |
| Vaccination during pandemic |  | Completed vaccination at 5 months of age(incl HAV, HBV) rate dropped by ca -17% , no change in HBV birth doses [117] | Reduction of 6.7% followed by increase.[116] | - 19.4% (in administered doses)[123] | Sustained coverage [113] |
| New diagnoses | -95% to -71% (HBV, HCV)[83] |  | No significant change (HAV) [114], decrease (HCV)[82] |  | -14% (HAV), no significant decline (HBV)[113] |

The choice of the endpoints is motivated by the findings of the modelling studies. For full results, see S1 Tables 7-9(TB), 12-13(viral hepatitis), 19-20(HIV). AFR = African Region, AMR=Region of Americas, EUR=European Region, SEAR=South-East Asian Region, WPR=Western Pacific Region, OR=odds ratio, IQR =inter-quartile range, HAV = hepatitis A, HBV = hepatitis B, HCV = hepatitis C
<sup>a</sup> % change w.r.t. control period, <sup>b</sup> difference w.r.t. control period

**Table 3.** Brief overview of observed impacts of COVID-19 on selected endpoints for malaria.

|  | WHO region |  |  |  |  |
| --- | --- | --- | --- | --- | --- |
|  | AFR | AMR | EMR | SEAR | WPR |
| **Malaria** |  |  |  |  |  |
| Diagnoses and treatment (Disruption: 5-50%) | 16 of 27 countries[52] | 8 of 16 countries [52] | 4 of 6 countries [52] | 5 of 8 countries [52] | 4 of 7 countries [52] |
| Cases – change attributed to COVID-19 | -40%[124], -50%[131] | Steady decrease[132] |  | +17%[126] | Decrease[130] |
| ITN campaigns | 34.3 of 122.2 million ITNs distributed (28%, end of August)[129] | Reduction of 60% in malaria-specific interventions[132] |  | Delay [126], higher distribution[128] |  |

The choice of the endpoints is motivated by the modelling studies. For full results, see S1 Tables 15-16.
AFR = African Region, AMR=Region of Americas, EMR=Eastern-Mediterranean Region, SEAR=South-East Asian Region, WPR=Western Pacific Region

### 3.1 Tuberculosis

Early on, modelling studies sounded alarm, highlighting that under-diagnosis of TB due to hindered access to care, reduced care seeking behaviour, and reduced health service and testing capacity are the greatest risk factors. The disruptions to services outweigh reduced transmission due to social distancing measures [30], which results in a pool of undetected and untreated TB with a prolonged duration of infectiousness leading to a delayed and prolonged rise in incidence and deaths. One model projected an excess 4.702.300 (8%) cases and 1044800 (12.3%) deaths [27] for 2020-25 and of 6.331.100 (10.7%) cases assuming 10% instead of 50% reduction in transmission [32]. Relating missed cases to deaths, a decrease in detection by 25% over three months increases TB deaths worldwide by 13% (1.660.000), a 50% decrease by 26% [28]. Other estimates predict 8-20% excess mortality [30, 31].

Mitigating measures included reducing the frequency of visits, using technology for remote consultations and restructuring directly-observed therapy schedules to provide longer medication supplies [111].

Nevertheless, TB notifications dropped worldwide following the introduction of lockdowns (**Fig. 3**, S1 Fig. 5).

**Fig. 3.**
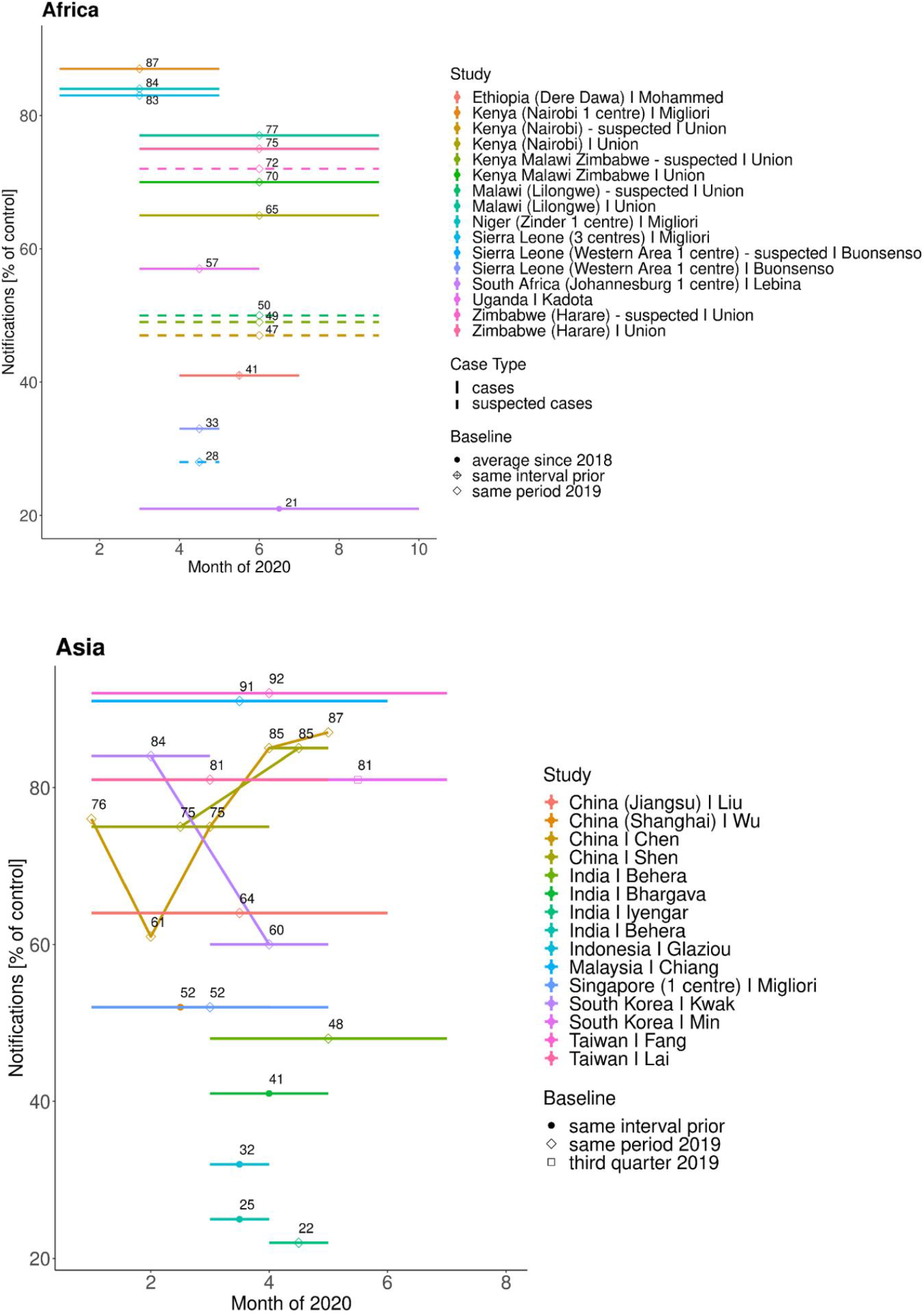
TB notifications in Africa and Asia. Overview of notifications over certain periods in 2020 (depicted by the horizontal lines) as percentage of the number of notifications in a control period (referred to as Baseline in the legend).

Preliminary data of notifications for the first six months of 2020 by 14 high burden countries shows a significant drop in notifications for all countries [111].

Reports from Sub-Saharan Africa show a trough in April 2020 with sustained reductions months after. Data from Uganda is used to predict 14% excess mortality [29].

India saw a 78% decrease in April with a slow recovery [102], partly attributed to staff deployment and delayed reporting. Data has been used to predict 19.6% (87711) additional deaths [26]. China saw the trough in February and entered the mitigation phase in April [100]. The impact on notifications in South Korea, Taiwan and Japan was less severe due to the predominant prevalence of latent TB [104].

Data from Europe shows a cumulated average reduction of 11% for Russia and 20% for Western Europe [109].

In the Americas, Paraguay reports a significant and sustained reduction from April [90] and seven centres in three countries a cumulative drop of 71% in April [109]. First results of effects on clinical outcomes show moderate effects on treatment completion, drug resistance screening and mortality varying by region. They are summarised in S1 Table 8.

The collateral impact of pandemic measures on TB services manifested immediately. In a WHO survey [111] the most common impediments were reallocation of staff, funding and testing capacity as well as reduced capacity/closing of facilities. More than half of Centers for Disease Control and Prevention-funded TB programs reported a partial/high impact to all items in a survey in April (staffing capacity, essential services, contact tracing, case reporting, etc.) [85]. In China [87], 3 out of 4 hospital beds for TB were closed during the pandemic phase. Moreover, lockdowns disrupted patient and staff access to services [62, 88]. Combined with fears of stigma and infection in the health care setting, visits to TB services plummeted. More than 50% of WHO programmes (28/30 high burden) [111] and 84% of 33 centres worldwide reported fewer TB outpatient visits [109]. Thirteen centres in China saw a reduction by 34% [87].

### 3.2 Viral hepatitis

Modelling studies regarding viral hepatitis examined the impact of treatment delays and vaccination interruptions. A delay in treatment for HCV under different scenarios led to significant excess mortality for e.g. liver cancer, depending on the duration of the program delay [43]. Another study [42], considering both direct and indirect impact of COVID-19, estimated increases in excess mortality for incident and prevalent liver cancer at various levels of infected population shares and using different relative risk (RR) scenarios. Compared to other considered cancers, increases in liver cancer deaths were at medium, and they increased with population infection rate and assumed RR. Excess deaths were also reported from a model-based risk-benefit analysis that compared vaccine continuation to vaccine interruption during the pandemic [44]. See also S1 Table 11. Several surveys attempted to measure the actual impact of the pandemic on service provision. The respondents on the provider side reported a decrease in testing volume, in consultations with diagnosed hepatitis patients and in numbers of (newly) treated patients for the considered period of 2020 compared to pre-pandemic times. These reductions were attributed to resource constraints (human or financial), and to cancellations of visits by patients (e.g. over 50% of cancelations were induced by patients in [82]). Most services were less impacted when regulations were less strict (e.g. in June, July, August 2020 [68, 75, 82]). Regarding changes in general care for chronic hepatitis patients, 80% of centres for chronic HCV patients in Germany did not see an impact [82], but low income countries partly faced limitations in treatment supply, mostly due to restrictions in transportation [83].

Findings from studies using other sources like notifiable disease records [113, 114] or vaccine records [116, 117] also varied. Some reported no significant declines in incident cases (e.g. [114] for HAV, or [113] for HBV), whereas others saw a decrease in notified cases in 2020 compared to pre-years, with the months of intensified social distance measures having the most reductions in reported cases [113] (for HAV). A decrease in vaccinations (routine childhood, including HBV as a hexavalent vaccine) was also seen [116, 117, 123].

Decreases in clinic visits and hepatitis care were reported throughout February to April 2020[83, 122], but the results varied by country [122]. Significant declines in HCV patients who kept their scheduled hospital visits in 2020 (March through May 2020 versus 2019) were reported[121]. Specifically for liver related morbidities, fewer hepatocellular carcinoma (HCC) patients were seen in 2020 and significant decreases for both HCC patients and treatment of active HCC patients were registered in early lockdown [112]. Treatment delay was longer in 2020, resulting in significantly higher severity (e.g. larger tumour size during the first diagnosis of HCC or worse cirrhosis severity score) [112, 119]. The period 2020 was an independent predictor for treatment delay above one month and for change/cancellation of treatment (multivariate analysis, OR 9.66; 95% CI: 2.85-32.72) [112]. On the contrary, other authors [120] found no significant difference in admissions or severity when comparing 2020 (March, April) to previous years for decompensated cirrhosis. A shorter length of hospital stays in 2020 showed an association with increased hospital readmission in 2020[120]. Although mostly anecdotal and not data-driven, statements from grey literature (S1 section 4) reported challenges for maintaining essential hepatitis services, such as effects on specimen transportation, testing and prevention efforts, re-purposed staff and suspension of routine clinic services. Concerns from European countries were raised on missed cases of acute HCV and delayed diagnoses of HCC in hepatitis patients.

### 3.3 HIV

Modelling studies identified ART interruption/decrease in viral load suppression as one of the leading individual factors increasing the 1-year HIV incidence by 6-29% (or even 126% [36]) at a 50% reduction for 3 to 6 months. Suspension of HIV testing (at 50%, 3 or 6 months) was responsible for about a 1% increase in the incidence, and the effect of fewer ART initiations was of a similar order. Regarding mortality, the leading factor was the same (ART interruption/decrease in viral suppression), causing an increase in 1-year mortality by 39%-87% at a 50% reduction (for 3 or 6 months). (See also S1 Table 22.)

In surveys and observational studies, a variety of outcomes addressed the impact of COVID-19 on retention in care. A lower ART adherence (self-reported, electronically recorded) during the pandemic was detected [53, 60], but also its immediate increase after the introduction of protective actions was noted [61]. In surveys, up to 15% of respondents assessed their ability to adhere as decreased (**Fig. 4**). About 20% of respondents (global, USA, Kenya) reported having difficulties keeping their HIV appointments or losing access to their health provider [57, 70, 72, 73]. In Uganda, 76% noted an impact of the restrictions on their ability to come to the clinic [65]. As of May-June 2020, 31% of 101 countries reported partial disruption to ART services (5%-50% of patients not treated as usual), 1% experienced severe disruption (>50% patients not treated as usual)[79].

**Fig. 4.**
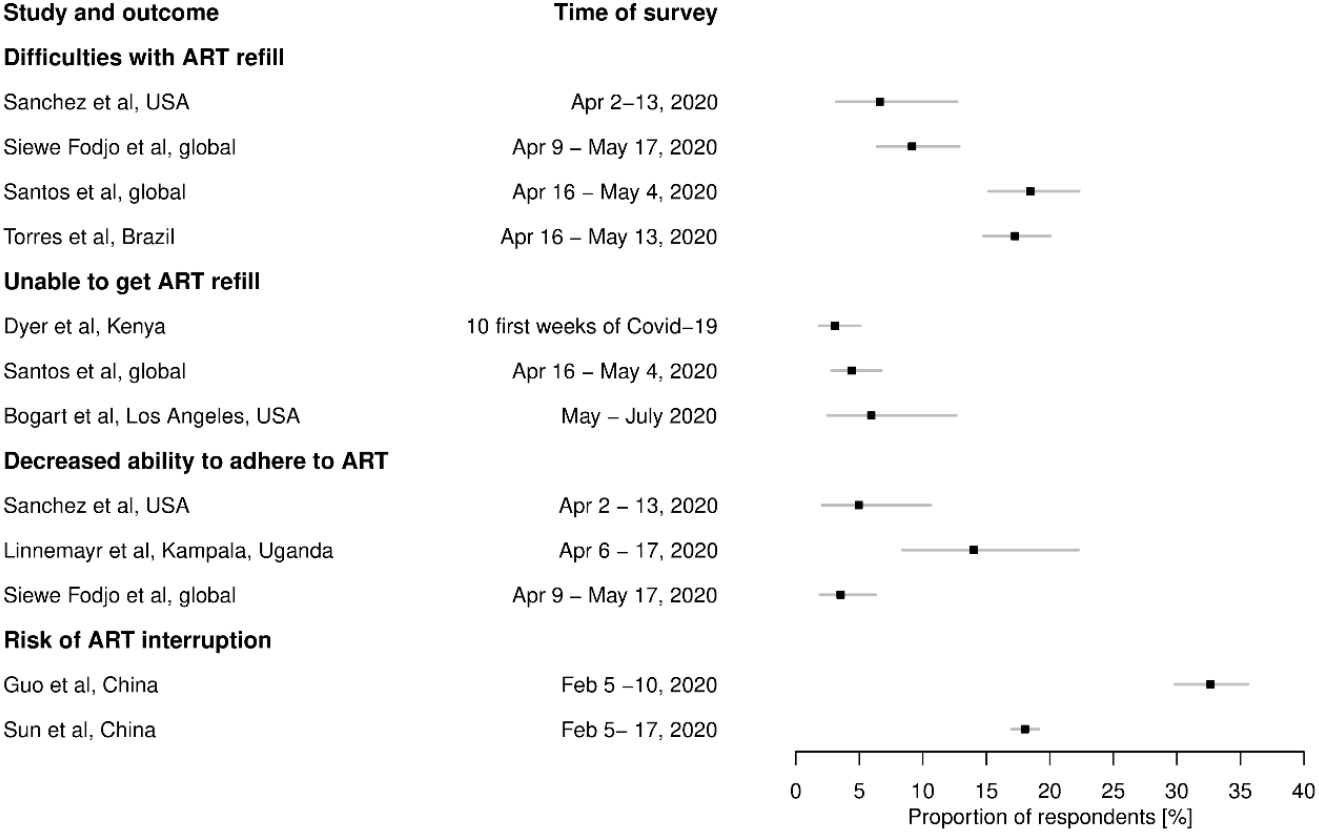
Self reported negative impacts of Covid-19 on ART refill and adherence. Reported are percentages with 95% confidence intervals based on Agresti-Coull method for binomial proportions. Note that the difficulties in refill may include also the inability to refill. The risk of ART interruption was defined as having medication for less than 10 days and not having a clear way how to refill by Sun et al [77], whereas Guo et al [58] considered only not having enough medication under the current traffic and movement restrictions. The majority of respondents in Siewe Fodjo et al [74] were from Belgium, Brazil and Eastern Europe, in Santos et al[73] from Brazil, France, Mexico, Taiwan and Russia. The numbers agree with 7% claiming no access and 21% limited access to ART in a global survey conducted in April-May 2020 based on about 2300 LGBT+ respondents[64].

Using health records to measure retention in care yielded mixed results: increase in missed visits and decrease in ART dispensation [153], no changes [157] partly thanks to adjustments in operational practices [150], or even an increase in ART consultations in April 2020 [160]. There were moderate decreases in viral load tests (a proxy for monitoring ART) in Croatia [133] and South Africa [147]. Further, 16/25 countries reported no declines in the number of people currently on ART [162]. Only very few (three) studies reported on the changes in the viral load of their patients. In one centre, 95.6% of 506 patients stayed aviraemic(Italy) [140]. In another, an adjusted odds ratio of 1.31 (95% CI 1.08 – 1.53) for unsuppressed viral load during the pandemic was reported (1766 patients, USA)[157]. Information on ART initiations was sparse. UNAIDS reported 75% (21/28) of the examined countries to „experience more sustained disruptions“ [162]. However, the reported changes ranged from -45% (Sierra Leone) to +88% (Nigeria) over March-August 2020. China saw a reduction of 34%[155], Uganda of 60% early in April, and of 37% later in June[41]. A moderate decrease in CD4+ tests (a proxy for ART initiations) was registered in South Africa [147].

The frequent initial decrease in facility-based HIV tests (by 30-90% in April-May 2020) was followed by a prospect of a rebound (see S1 Fig. 6). In surveys, providers claimed a decreased demand for and a decreased ability to test [56] (USA), as well as partly severe reductions in testing volumes [75] (European region). About 20-30% of MSM respondents in the USA stated being prevented from getting tested [76], or experienced difficulties getting tested when tried [67, 72].

The numbers of new HIV diagnoses on national/regional level decreased early (by 12%[139], 23%[134], 24%[155] and 45% [133]). Single centres or programmes reported both a decrease [41, 66, 135, 137, 153] and an increase[151], some noted an increased proportion of late presenters [139, 151]. An increase in the positivity rate was noted in Japan and in collated data from Malawi, Zimbabwe, and Kenya (S1 Fig. 6).

Details on further outcomes as listed in **Fig. 2** are given in S1 section 10.

### 3.4 Malaria

The modelling studies uniformly reported an increase in malaria cases and malaria-associated deaths and morbidity. In the worst-case scenario (WHO scenario 9: no insecticide-treated nets (ITN) campaign, continuous distribution of ITNs reduced by 75%, treatment with effective malaria medication reduced by 75%), an increase in malaria cases of 23% and malaria-associated deaths of 102% was predicted in sub-Saharan Africa [49]. Another model predicted an increase of 113% (165%) in malaria cases (deaths) [45] when malaria prevention measures (ITN campaigns, seasonal malaria chemoprevention (SCM) and treatment of clinical malaria cases) were interrupted over 12 months. The increase in malaria-associated deaths by 2024 could be as high as 36% in high burden countries [31]. The single intervention with the greatest impact was either the distribution of ITNs [31, 45] or access to effective antimalarial treatment [41, 46, 49]. If planned ITN campaigns were to be cancelled in sub-Saharan Africa due to the COVID-19 pandemic, in the worst-case scenario (WHO Scenario 9) the increase in malaria cases (+ 41.9 million) and malaria-associated deaths (+ 331 630) could be almost twice as high as in countries where campaigns were still being implemented or had already taken place in 2019 [49]. The impact of COVID-19-related reduction of health service coverage and, therefore, reduction in malaria-associated prevention measures on the mortality of mother and child was predicted to be 1-2% per individual intervention considered, depending on the severity of the interruption [47].

We note in passing that whereas all the modelling studies associate anti-pandemic measures with interruption of malaria prevention services, in [48] ITN distribution is inversely proportional to the number of COVID-19 infections. This means that implementing more anti-pandemic measures, which implies a decrease in COVID-19 cases, results in more ITNs distributed and thus in a lower negative impact on malaria transmissibility as measured through vectorial capacity. Unfortunately, the authors did not discuss the assumptions made and seemed to ignore the possibly longer delay between declining COVID-19 cases and lifting of the anti-pandemic restrictions that affect the ITN distribution. The predictions of the modelling studies may be compared to numbers reported by observational studies, whose findings are rather mixed (S1 Tables 15-16).

Regarding the actual impact of COVID-19, several studies [50-52, 126, 129, 131, 132] reported logistical problems and supply bottlenecks, which led to delays or interruptions of malaria prevention measures. It was also observed that delayed delivery of ITNs can lead to an increase in malaria cases, especially those transmitted locally, by increasing the potential for malaria transmission [126, 131]. The deployment of staff to contain the COVID-19 pandemic is likely to compromise malaria surveillance [51, 126, 131]. On the other hand, fear of infection with COVID-19 leads to reduced use of health services and a consequent drop in the number of malaria cases [124, 130, 132]. In addition, access to and supply of health services may be reduced due to the pandemic [50-52, 131, 132]. By developing action plans and stockpiling sufficient material, malaria prevention can continue during a pandemic and the number of malaria cases can be reduced [127, 128]. However, the adaptation of prevention measures to the conditions and restrictions of the COVID-19 pandemic, e.g., distribution of ITNs from door-to-door, required an increased expenditure of time and money [49, 51, 131]. Hygiene concepts were elaborated and implemented, the provision of personal protective equipment and staff training caused additional effort [125, 128-130].

## 4. Discussion

Quantified information on the COVID-19 impact on the treatment and prevention of TB, viral hepatitis, HIV/AIDS and malaria is important when assessing the impact of the pandemic on the eradication of these diseases. In this respect, modelling studies play a crucial role since the effects on new cases and deaths occur only with a delay. The modelling studies collected in our systematic review played yet another role. Early in the pandemic, they contributed to the awareness of the collateral impact of the pandemic and prompted a re-shift towards maintaining essential services. They were the basis for official recommendations for adjustments in service and treatment provision (see, e.g., [165-167] for HIV, [168] for TB, [169] and country action plans [51, 52, 131, 169] for malaria). In summary, for TB, the modelling studies concluded that the reduction in droplet transmission through pandemic measures is outweighed by the disruptions to services, and the ensuing under-diagnosis results in a larger pool of undetected and untreated (thus remaining infectious) cases, leading to a delayed but prolonged rise in incidences and deaths. Some countries implemented active case finding programs to “catch-up” on pandemic under-detection [25]. Early modelling studies for HIV, e.g. [36], warned about serious consequences on both HIV incidence and HIV-related mortality if ART interruption and subsequent decrease in viral load suppression should occur. The modelling studies regarding malaria identified ITN campaigns and maintaining access to effective antimalarial drug therapy as the most influential individual prevention measures. However, three publications [41, 46, 49] used the same model assumptions, so that the same qualitative findings are not surprising. Hepatitis modelling predicted excess mortality due to delayed HCV treatment, or interruptions in childhood vaccination.

However, seen as predictions of future real-world impacts, many of the modelling scenarios seem inadequate in retrospect. They were based on the shock observations of uncoordinated early lockdowns and could foresee neither the prolonged and recurrent restrictions nor adaptations towards a “normality” under pandemic conditions. The parameters were often based on expert consultation rather than real-world data. Under the interruption of malaria prevention measures, the modelling studies predicted dramatic increases in malaria cases and malaria-associated deaths. These were, however, not reported with such clarity in the observational studies. One reason may be the short observational periods of 2 to 9 months. Another reason, however, could be a successful adaptation of the prevention programs to the anti-pandemic measures. Updated modelling by UNAIDS [162] for HIV concludes that the effects of the pandemic on HIV incidence and mortality are restricted to a short horizon not stretching beyond 2024, so the 2030 goals for ending the AIDS pandemic can still be reached [170]. Nevertheless, an important assumption in this modelling is an efficient global vaccination.

Our systematic review aimed at mapping the actual extent of service and treatment disruptions due to COVID-19. For all four diseases assessed, we found a large heterogeneity of the effects both between and within countries. This reflects not only the strictness of the implemented anti-pandemic measures, but also the pre-pandemic organization of the services, the degree of difficulties connected with a change to digital technologies and the degree of success in adjusting the service provision to the pandemic restrictions. The heterogeneity is reflected in the observational studies, reports of clinical outcomes, as well as surveys.

For example, clinical outcomes of TB suffered particularly where services include a larger share of outreach [89], where access to services was restricted and a change to digital technologies harder to implement. In particular, outcomes in South Korea [95] compare favourably to China [87, 97], where services suffered a higher degree of restructuring, and were better in Malawi than Kenya and Zimbabwe, as Malawi did not impose a lockdown and maintained availability and accessibility. Another example is service provision. Besides many studies stating a negative effect, some centres, programs or countries reported no change or even positive changes in particular indicators (e.g. no decrease in HIV testing volumes in Rwanda [162], or a successful continuation of malaria prevention despite the challenges [127, 128]). Similarly, for viral hepatitis, both a decrease and an increase in services from different centres within one country was reported [68] and differences were seen also in the same area of care (e.g. regarding the primarily treated viral hepatitis disease (HBV or HCV), direct-acting antivirals for HCV decreased [68, 83], but testing sometimes increased [68]).

The negative impact on service provision differed in causes across the four diseases. For malaria, transportation and supply bottlenecks were a challenge for the main prevention measure, the distribution of ITNs. TB services suffered primarily from the redirection of resources. TB staff was already skilled in handling infectious patients, infection control protocols, contact tracing and isolations. Furthermore, GeneXpert machines were reassigned to SARS-CoV-2 diagnosis where laboratory capacities were limited. Besides re-allocation of staff, viral hepatitis and HIV services suffered from a reduction in face-to-face counselling as a consequence of applying physical distancing measures requiring, e.g., limitation of capacity [68, 75].

The fear of acquiring SARS-CoV-2 in hospital settings was a near-universal contributor to a decline in service usage and healthcare-seeking behaviour, with 50% of hepatitis appointments cancelled by patients [103]. Additionally, for TB, due to the overlap of symptoms, this was aggravated by the fear of being diagnosed with SARS-CoV-2 instead of TB, which in many countries carried a heavy stigma. Nevertheless, data on suspected cases from Africa [99, 111] shows that health-seeking behaviour was impacted more for less severe cases and that inequalities are reflected in the data [32] showing a greater reduction in attendance for children than adults, and for women than men. Regarding malaria, some local reports identified the fear of SARS-CoV-2 as problematic and described active counter-measures [125, 130]. In the case of HIV, restrictions in transportation and movement were another reason impeding service usage in some areas [58, 65]. For viral hepatitis, the heterogeneity in findings was also linked to the origin of survey participants and centre location [122]. Replies from low-income countries particularly alluded to people identified with viral hepatitis not being referred to care or further medically investigated, or that treatment shortages had led to interrupted care [83]. Studies reporting on certain indicators at several time points throughout the first half-year of 2020 enable a glimpse into the regional dynamics of the pandemic. In the case of reduced notifications and the implied under-detection of TB [32], data from the first wave in China show a direct relation to the control measures and degree of restructuring. The troughs coincide with the lockdowns, and the data suggest that the recovery phase may be slow and delayed even for a short and contained crisis phase [100]. In the case of HIV, data on post-exposure-prophylaxis prescriptions from Spain [154] and Australia [136] suggest that the recovery from an initial decrease started already before the lockdown measures were relieved. Thus, the deep initial decrease may be unique to the first wave of the pandemic and reflect an over-reaction to the unprecedented situation. A different reaction to the pandemic was an increase (approximately. 20%) in HIV related clinic visits in KwaZulu-Natal immediately after the introduction of lockdown [156]. This may hypothetically reflect the urgency in medication refill before anticipated restrictions to services.

Despite the valuable insights, the published evidence has several limitations.

For example, although the surveys on COVID-19 impact on viral hepatitis provide information on alternative offers (e.g. online consultations) made for the patients, they are often based on “best estimates” rather than numerical information, which is a limitation when assessing the overall impact of the pandemic. Studies using hepatitis health records did not conduct causality assessments. When evaluating vaccine coverage, no adjustment for eligible vaccine recipients was conducted, although, vaccinations might be influenced by birth rates and general trends in vaccination (decreases;[116]). Significance in changes was rarely assessed, and only absolute numbers or %-change compared to pre-pandemic periods was reported [114]. Some studies had restrictive inclusion criteria (only those with sustained virological response among HCV patients [121]), which may limit conclusions for hepatitis patients.

Similarly, although the changes in HIV tests volumes were based on the highest number of studies relying on health records data, only a few studies looked at longer historical records of testing volumes and judged the observed numbers in 2020 against the historical background accounting for long-term trends and the usual variation. Most studies used only a single time interval for the comparison. If the corresponding time interval in 2019 was used, one could hope that possible seasonality effects were accounted for; however, if the comparison was made to the pre-pandemic time in 2020, the results may be distorted by seasonality. Even though we may confidently attribute a drop of 40% and more to the pandemic situation, without gauging the usual variability, we cannot be sure whether lower drops of 20% or 10% are still real drops.

Finally, the collected published evidence in this review covers only the first half-year of the pandemic. Thus, the published numbers do not show how the situation evolved after June or August 2020. The observation time is often too short to assess important mid- to long-term outcomes, such as the retention in care or the viral load suppression of HIV patients, as well as long-term care outcomes for HIV, TB or hepatitis, or changes in malaria cases and malaria-related deaths.

## 5. Conclusions

The evidence collected in this review suggests an apparent recovery from the first pandemic shock, according to several indicators. However, it does not show whether this recovery has been stable, neither do we know whether the negative impacts were avoided during the second and third COVID-19 waves. Further, it would be of interest to see if and in which form the adaptations in services, e.g. [171-174], have been kept and what their long-term influence, for example, on the retention in care is.

Projections of the long-term impact of sudden events using data only from a short-term intensive phase and setting parameters based on expert opinion should be treated with caution. Given the recurring waves and lasting impact of the pandemic, with many countries in HIV, TB or malaria-endemic regions not expecting significant immunization until 2023, and with the mid- to long-term effects of adaptation and normalisation unknown, the real consequences for the fight against leading infectious diseases worldwide will only manifest over the coming years.

## Supporting information

PRISMA 2020 checklist

## Data Availability

All data analysed in the present work are contained in the manuscript and the supplementary material.

## Author contributions

Conceptualization: BK, TH, JJO, JW, MH, BL

Data Curation: BK, TH, JJO, JW, MH, BL

Formal Analysis: BK, TH, JJO, JW, MH, BL

Funding Acquisition: BL, JJO

Investigation: BK, TH, JJO, JW, MH, BL

Methodology: BK, TH, JJO, JW, MH, BL

Project Administration: BL, BK

Software: BK, TH

Supervision: BL

Validation: BK, TH, JJO, JW, MH, BL

Visualization: BK, TH

Writing – Original Draft Preparation: BK, TH, JJO, JW, BL

Writing – Review&Editing: BK, TH, JJO, JW, MH, BL

## Data sharing

All data underlying results in this review are either presented directly in the main text or shown in S1, sections 7-10, in particular in the S1 Tables 7-22.

## Supporting information captions

### Text

S1 section 1: Search strategy in databases

S1 section 2: Search strategy for grey literature

S1 section 3: Post-hoc amendments to the study protocol

S1 section 4: Details of the screening process and the “near-misses” S1 section 5: Risk-of-bias assessment tools

S1 section 6: Risk-of-bias assessment results S1 section 7: Tuberculosis

S1 section 8: Viral hepatitis S1 section 9: Malaria

S1 section 10: HIV

### Tables

S1 Table 1: Details of the grey literature search

S1 Table 2: Details on grey literature findings regarding viral hepatitis

S1 Table 3: Risk-of-bias tool for modelling studies

S1 Table 4: Risk-of-bias tool for surveys

S1 Table 5: Risk-of-bias tool for quantitative studies

S1 Table 6: Overview of studies related to TB with respect to the outcome and type of evidence/data source

S1 Table 7: Overview of findings describing the impact of COVID-19 on TB notifications

S1 Table 8: Overview of findings regarding indirect impact of COVID-19 on different clinical outcomes related to TB

S1 Table 9: Overview of published evidence regarding impact of COVID-19 on TB-related service provision

S1 Table 10: Overview of the modelling studies regarding indirect impact of COVID-19 on TB

S1 Table 11: Findings of the modelling studies related to indirect impact of COVID-19 on viral hepatitis

S1 Table 12: Findings of the surveys related to indirect impact of COVID-19 on viral hepatitis S1 table 13: Findings of studies relying on hospital or notifiable disease records, registries and databases, when assessing the indirect impact of COVID-19 on viral hepatitis

S1 Table 14: Overview of studies related to malaria with respect to various characteristics

S1 Table 15: Overview of projected and observed impact of COVID-19 on malaria cases. S1 Table 16: Overview of projected and observed impact of COVID-19 on malaria deaths

S1 Table 17: Projected consequences of interruption of malaria prevention measures on malaria cases and deaths and overview of observed disruptions of the measures

S1 Table 18: Overview of studies related to HIV with respect to various characteristics

S1 Table 19: Self-reported HIV service interruption

S1 Table 20: Overview of studies reporting on ART retention and engagement in care

S1 Table 21: Overview of findings related to HIV testing and new diagnoses

S1 Table 22: Overview of the impact of individual HIV service/care interruptions on HIV-incidence and mortality as reported by the modelling studies

S2 Table 1: PRISMA checklist

### Figures

S1 Fig. 1: Risk-of-bias assessment for surveys related to viral hepatitis, tuberculosis and malaria

S1 Fig. 2: Risk-of-bias assessment for surveys related to HIV

S1 Fig. 3: Risk-of-bias assessment of quantitative studies related to viral hepatitis and tuberculosis

S1 Fig. 4: Risk-of-bias assessment of quantitative studies related to malaria and HIV

S1 Fig. 5: TB notifications in Americas and Europe

S1 Fig. 6: Reported reduction in HIV testing volumes and changes in the positivity rate

S1 Fig. 7: Impact on PrEP usage

S1 Fig. 8: Reduction in PEP prescriptions

