## Supplementary file S1 for "Impact of COVID-19 pandemic and anti-pandemic measures on tuberculosis, viral hepatitis, HIV/AIDS and malaria – a systematic review"

### 1. Search strategy in databases

**Database: PubMed**, https://pubmed.ncbi.nlm.nih.gov/advanced/
Search 28.09.2020 and 13.01.2021:
((COVID-19) OR (SARS-Cov-2)) AND ((hepatitis) OR (tuberculosis) OR (HIV) OR (malaria))

("severe acute respiratory syndrome coronavirus 2"[Supplementary Concept] OR "severe acute respiratory syndrome coronavirus 2"[All Fields] OR "ncov"[All Fields] OR "2019 ncov"[All Fields] OR "covid 19"[All Fields] OR "sars cov 2"[All Fields] OR (("coronavirus"[All Fields] OR "cov"[All Fields]) AND 2019/11/01:3000/12/31[Date - Publication]) OR ("severe acute respiratory syndrome coronavirus 2"[Supplementary Concept] OR "severe acute respiratory syndrome coronavirus 2"[All Fields] OR "sars cov 2"[All Fields])) AND ("hepatitis"[MeSH Terms] OR "hepatitis"[All Fields] OR "hepatitides"[All Fields] OR "hepatitis a"[MeSH Terms] OR "hepatitis a"[All Fields] OR ("tuberculosi"[All Fields] OR "tuberculosis"[MeSH Terms] OR "tuberculosis"[All Fields] OR "tuberculoses"[All Fields] OR "tuberculosis s"[All Fields]) OR ("hiv"[MeSH Terms] OR "hiv"[All Fields]) OR ("malaria"[MeSH Terms] OR "malaria"[All Fields] OR "malarias"[All Fields] OR "malaria s"[All Fields] OR "malariae"[All Fields]))

Translations

COVID-19: "severe acute respiratory syndrome coronavirus 2"[Supplementary Concept] OR "severe acute respiratory syndrome coronavirus 2"[All Fields] OR "ncov"[All Fields] OR "2019-nCoV"[All Fields] OR "COVID-19"[All Fields] OR "SARS-CoV-2"[All Fields] OR ((coronavirus[All Fields] OR "cov"[All Fields]) AND 2019/11:3000[pdat])

SARS-Cov-2: "severe acute respiratory syndrome coronavirus 2"[Supplementary Concept] OR "severe acute respiratory syndrome coronavirus 2"[All Fields] OR "sars cov 2"[All Fields]

hepatitis: "hepatitis"[MeSH Terms] OR "hepatitis"[All Fields] OR "hepatitides"[All Fields] OR "hepatitis a"[MeSH Terms] OR "hepatitis a"[All Fields]

tuberculosis: "tuberculosi"[All Fields] OR "tuberculosis"[MeSH Terms] OR "tuberculosis"[All Fields] OR "tuberculoses"[All Fields] OR "tuberculosis's"[All Fields]

HIV: "hiv"[MeSH Terms] OR "hiv"[All Fields]

malaria: "malaria"[MeSH Terms] OR "malaria"[All Fields] OR "malarias"[All Fields] OR "malaria's"[All Fields] OR "malariae"[All Fields]

Search 13.01.2021: publication date from 2020/09/29

**Database: preView** (includes medRxiv, bioRxiv, ChemRxiv, ResearchSquare, arXiv, Preprints.org), https://preview.zbmed.de
Search 28.09.2020: tuberculosis OR HIV OR malaria OR hepatitis
Search 13.01.2021: tuberculosis OR HIV OR malaria OR hepatitis with
Publication date 2020/09/29
Publication date 2020/09/30
Publication date 2020/10
Publication date 2020/11
Publication date 2020/12
Publication date 2021/01

**Database: Scopus**, https://www.scopus.com/search/form.uri?display=advanced
Search 28.09.2020: TITLE-ABS-KEY( ("SARS-CoV-2" OR "COVID-19") AND ("tuberculosis" OR "HIV" OR "hepatitis" OR "malaria"))
Search 13.01.2021: TITLE-ABS-KEY( ("SARS-CoV-2" OR "COVID-19") AND ("tuberculosis" OR "HIV" OR "hepatitis" OR "malaria")) AND PUBDATETXT(„29 September 2020" or „30 September 2020" or „October 2020" or „November 2020" or „December 2020" or „January 2021")

Division of the identified records into groups according to TB/Malaria/Hepatitis/HIV: relevant papers for each disease were found using the search function in EndNote X9 and the following search terms in any field:
TB: tuberculosis OR tuberculoses OR TB

HIV: HIV OR HIV-1
Hepatitis: hepatitis OR hepatides OR HBV
Malaria: malaria OR malariae OR malarias

### 2. Search strategy for grey literature

**S1 Table 1 Details of the grey literature search.**

| Disease | Date of the search | Websites searched |
|---|---|---|
| Viral hepatitis | Sep 30, 2020<br>Jan 28, 2021 | The main medical societies in the field:<br>• The European Association for the Study of the Liver (EASL, www.easl.eu)<br>• The American Association for the Study of Liver Diseases (AASLD, www.aasld.org)<br>The largest umbrella-NGO in the area of hepatitis<br>• The World Hepatitis Alliance (WHA, www.worldhepatitisalliance.org)<br>Platform for international hepatitis news<br>• Hepatitis Voices(www.HepVoices.org) |
| Tuberculosis | Dec 2020 | WHO (www.who.int), TB Union (theunion.org), The Global Fund (www.theglobalfund.org) |
| HIV/AIDS | Feb 1, 2021 | WHO (www.who.int), UNAIDS (www.unaids.org), International AIDS Society (www.iasociety.org), Kaiser Family Foundation (www.kkf.org), European AIDS Treatment Group (www.eatg.org), Sensoa (www.sensoa.be), PEPFAR (www.state.gov/pepfar), The Global Fund (www.theglobalfund.org), AMPATH (www.amptahkenya.org), CAPRISA (www.caprisa.org), the virtual AIDS 2020 conference (http://programme.aids2020.org/) |
| Malaria | Jan 30, 2021 | WHO (www.who.int), RBM Partnership to End Malaria (https://endmalaria.org/),The Global Fund (https://www.theglobalfund.org/en/) |

### 3. Post-hoc amendments to the study protocol

For surveys, we accepted an implicit comparator, i.e. questions with wording "due to COVID-19", or "under current travel restrictions".

In the case of HIV, we adopted the following two additional exclusion criteria:

- small sample size: 20 or fewer respondents if not speaking on behalf of programmes or organizations,
- surveys in which a certain group of people was asked about the COVID-19 impact on another group (e.g. health care professionals were asked about the impact on patients).

We also did not consider survey questions regarding anticipated difficulties due to COVID-19, as opposed to experienced difficulties.

### 4. Details of the screening process and the "near-misses"

#### Tuberculosis

Three publications classified as near misses in the full-text screening. Two did not contain data related to COVID-19 [1, 2], one did not provide a comparator for the absolute impact [3].

#### HIV/AIDS

In full-text screening, we excluded 14 full-texts under the label: insufficient reporting/data. Here we give their details. Three [4-6] did not contain any precise quantifications in their statements. They reported endangered ART supplies in Indonesia[4], maintaining HIV therapy among people with substance use disorder in Ukraine[6] and cessation of community HIV testing in Birmingham, Alabama[5]. Four publications [7-10] did not contain information to which period exactly the reported data refer. They reported a decrease in PEP prescriptions (China)[8], interruptions in medication uptake(China)[9], no interruption in outpatient services and therapy withdrawal(Italy)[7] and self-reported problems to access HIV medication by 4% of respondents (Argentina)[10]. Further, four publications were based on surveys of and about individuals with 20 and fewer respondents, reporting no interruptions to HIV care(USA) [11] or no interruptions to access to ART(USA) [12], a decrease in PrEP usage(USA) [13] and reasons preventing the uptake of facility-based HIV testing(Uganda)[14]. Further, two publications did not provide a clear comparison of their observed numbers of rescheduled/missed visits to pre-pandemic times (USA, Rwanda) [15, 16]. Finally, one publication did not describe the comparator for the reported reductions in HIV tests (-59%), VMMC (-99%), ART initiation (-51%), viral load sample collection (-29%), citing a not accessible rapid assessment of Ministry of Health and Child Care in Zimbabwe [3].

Related to two outcomes: median visits per day and delivered ART doses, one publication[17] did not specify the comparator pre-lockdown period and reported a decrease in median visits per day during March 9 – June 10, 2020 ( 7 vs 13) and no change in the number of delivered ART doses (thanks to adaptations in care).

From the grey literature search, we count as near misses five conference abstracts not containing enough information on the outcome or periods concerned, so that the reported numbers cannot be put into perspective [18-22]. We neither included rapid assessments of the situation in HIV health care in Europe based on surveys of respondents affiliated with organizations advocating or serving PLHIV, i.e. not directly providing the services nor necessarily using them[23, 24]. Similarly, we did not include a survey mapping the situation from the viewpoint of NGOs and community-based groups serving PLHIV[25].

**Viral Hepatitis**

During the screening process, 581 and 193 records from 2020 and 2021 searches, respectively, were screened by title and abstract; 49 of them were screened in full text. The main reasons for exclusion were that studies focused on COVID-19 among hepatitis patients or on effects of COVID-19 on liver and associated diseases. Further, treatment guidelines were frequently published. Some economic calculations on future effects of medicinal products were excluded as well as genetic studies and investigations from entirely basic and laboratory science. By full-text screening, one study[26] was excluded due to insufficient reporting. The author mentions 375 patients with chronic active HBV registered at a hospital in Duhok city (Kurdistan region, Iraq) who did not receive the required care for the HBV infection during the lockdown. However, neither the duration of the lockdown nor the missing care is further specified.

Conducting the grey literature search, we found mostly webinars, toolkits for advocacy, patient associations and (patient) information material, position statements and guidance on clinical care for patients with (chronic) hepatitis, liver derangement or transplantations during COVID-19 (e.g. https://easl.eu/wp-content/uploads/2020/04/EASL-ESCMID-COVID-19-Position-Paper.pdf). A link announcing a survey among medical service providers and experts on the impacts of COVID-19 on clinical care or waiting-list management of transplant centres worldwide hinted at future resource of valuable data and evidence. A detailed overview of the found grey literature evidence is in S1 Table 2. Except for one, the items found were not included for data extraction due to their limited scientific expression (missing data, personal opinions, web seminars), or because the information included was part of published manuscripts already identified by our systematic search (e.g. the results in [27]). The statements from health care professionals, health policy-makers, and staff of WHO regional offices on experiences or status quo of care during the pandemic reported challenges for maintaining essential hepatitis services, such as effects on specimen transportation, testing and prevention efforts, re-purposed staff and suspension of routine clinic services. Concerns from European countries were raised on missed cases of acute HCV and delayed diagnoses of HCC in hepatitis patients.

**Malaria**

In the full-text screening, we identified only one publication [28] as a near miss. Its focus is not on evaluating the impacts of COVID-19, so there is no comparison to pre-covid times. The paper reports on digitalized mass ITN distribution in Benin, with the distribution phase being affected by COVID-19. This required quick adjustments, and the campaign ended successfully with 94.16% of the enumerated population receiving ITNs.

**S1 Table 2 Details on grey literature findings regarding viral hepatitis.** Note that coinfection with COVID-19 is in general of no interest for our review.

| Website | Details on the found materials and reasons for exclusion or indication of inclusion |
|---|---|
| www.easl.eu | Webinar of a discussion among hepatologists/experts on coping strategies with COVID-19, e.g., challenges for maintaining essential hepatitis services, among them effects on specimen transportation, testing and prevention efforts, re-purposed staff and suspension of routine clinics. Also, concerns from European countries on missed cases of acute HCV and delayed diagnoses of HCC in hepatitis patients are expressed, but no impact measure is stated.<br><br>Link to survey launched in September 2020 and sent to 470 transplant centres to collect and analyse regional practices on how liver transplant patients were affected by the pandemic - no results yet.<br><br>Cross-reference to a survey done by the WHA (see below). |

| | |
|---|---|
| | Toolkit for patient associations, e.g., on how to communicate COVID- information to patients and how to advocate for better treatment of liver disease patients (for media). EASL–ESCMID Position Statement on clinical care for patients, e.g., how healthcare providers can reduce transmission and prioritise patients, links to published research, covered by our systematic search (COVID risk and mortality among patients liver disease/viral hepatitis). – coinfection |
| www.aasld.org | Several Webinars on COVID-19 and liver disease (management, guidance, treatment, clinical insights and case studies); perspectives on impacts for hepatitis care and personal reports including impact on liver transplantation (US). – personal reports and experiences |
| www.worldhepatitisalliance.org | Webinars with statements from individuals on COVID-19 and the liver and link to a EASL/WHO discussion on the potential impact of COVID-19 on civil society-delivered hepatitis services and patients. – no evidence on the actual impact<br><br>Member survey on COVID-19 pandemic effects on hepatitis services and patients (results included in the extracted data); accessed 30th September, 2020<br><br>Medical advice for individuals living with hepatitis and being infected with COVID-19. – coinfection |
| www.HepVoices.org | News/information on community pharmacy that has postponed a HCV service from April 2020 to September 2020, but no impact measure stated: https://hepvoices.org/2020/08/hepatitis-c-service-delayed-by-covid-19-to-launch-on-september-1/, accessed 28th January 2021<br><br>Reference to a summary of statements received from some US health departments on experiences, concerns, requests, and strategies for STI care, incl. viral hepatitis. It was reported that programs have reduced or suspended services and activities in response to COVID-19 due to reduced staff. Also mentioned: suspension of outreach, education, and prevention; clinic closures, reductions in clinical services. https://www.naccho.org/blog/articles/lhd-hiv-sti-and-hepatitis-programs-respond-and-adapt-to-covid-19. Redirected from HepVoices.org, accessed 28th January 2021. – no numerical information<br><br>Reference to a press article on reduction in HBV-birth doses in India "19.4 per cent drop in Hepatitis-B birth doses administered and 31 per cent drop in vaccination sessions held in health facility and outreach sessions from April to June 2020 as compared to same period last year as per Health Management Information System (HMIS), in the light of the COVID-19 pandemic," https://hepvoices.org/2020/09/19-4-drop-in-hepatitis-b-birth-doses-31-drop-in-vaccination-sessions-in-april-june/, accessed 30th Sept 2020. – included |

### 5. Risk-of-bias assessment tools

We used three different tools for the risk of bias (RoB) assessment according to the type of study. For modelling studies, the tool is based on [29] and we added questions (inspired by [30]) assessing the reporting of the model and sources of data informing the model parameters (S1 Table 3). For surveys, we made a minor adjustment to the Risk of Bias Instrument for Cross-Sectional Surveys of Attitudes and Practices [31] based on ideas in [32, 33], see S1 Table 4. Finally, for other quantitative studies, we compiled the RoB tool from ideas in Appendices F, G in [34], covering two aspects of external and internal validity each, and including an assessment of possible selective reporting (S1 Table 5).

**S1 Table 3 Risk-of-bias tool for modelling studies.**

| **Modelling studies** | |
|---|---|
| 1.Does the model include real world data? | Yes/No |
| 2. Is the model thoroughly described, or a reference to such a description is given, and are parameter values of the model addressed? | Yes/No |
| 3. Is the model under discussion a dynamic model (transmission model)? | Yes/No |
| 4. Are uncertainty analyses conducted on key assumptions which could have impacted the conclusion? | Yes/No |
| 5. Is the source of the data informing the model based on official (routine) records (hospital, national)? (As opposed to expert opinion or convenience data.) | Yes/No |

| | |
|---|---|
| Overall RoB conclusion | No concerns: all „Yes“<br>Moderate concerns: „No“ only in point 2. and/or 5.<br>Major concerns: all other cases |

**S1 Table 4 Risk-of-bias tool for surveys.**

| **Surveys** | **LR= low risk, MR= medium risk, HR= high risk** |
|---|---|
| 1. Is the source population representative of the population of interest? | LR: random sample from a representative population roster (e.g. national registry or all centres in a country) or taking everyone in the population<br>MR: single centre/city/region or non-random sampling from a registry; random sample from participants in a different trial<br>HR: convenience sample, self-recruitment |
| 2. Is the response rate adequate? | LR: >75%<br>MR: ≤75% and respondents are similar to the target population (or to non-respondents) in important characteristics<br>HR: <40% and no characteristics are reported, or differences between respondents and non-respondents are reported<br>Not reported |
| 3. Is there little missing data within questionnaires? | LR: up to 10%, MR: up to 20%, HR: more than 20%, Not reported |
| 4. Was the questionnaire tested, or alternatively, are the exact questions reported? | LR: pilot testing + some clinimetric properties (e.g. face validity, construct validity, comprehensiveness) assessed in a similar population, or the exact question reported and seeming understandable, simple, addressing what it should<br>MR: only pilot testing, or clinimetric properties in a different population; or the question only rephrased<br>HR: no testing at all, or testing not reported, neither reporting of the exact questions |
| 5. Was the survey administered at a time, when the effect of the exposure could have already manifested? | LR: after 6 or more weeks of the exposure<br>MR: in week 2-6 of the exposure<br>HR: in the first 2 weeks of the exposure |

**S1 Table 5 Risk-of-bias tool for quantitative studies.**

| **Other quantitative studies** | **LR= low risk, MR= medium risk, HR= high risk** |
|---|---|
| 1. Are the population or source area and the data sources well described? | LR: good description of both the population variables and data origin<br>MR: limited description of the population variables and/or data origin<br>HR: insufficient description of the population and the data origin |
| 2. Is the eligible population or area representative? | LR: international, national. MR: regional, multi-centre, HR: local, single centre |
| 3. Are the control intervals and control cohorts appropriate? | LR: comparable cohorts and several previous intervals for comparison (capturing the usual variation in the outcome)<br>MR: comparable cohorts, but only a single interval chosen for comparison and its choice is justified<br>HR: non-comparable cohorts and/or a single comparison interval chosen without comments, or no specification of the comparison |
| 4. Is the observation interval meaningful? | LR: long enough for outcome to occur (clinical) OR data covering lockdown and after-lockdown periods<br>MR: data covering fully lockdown but no after-lockdown period<br>HR: not long enough for outcome to occur (clinical) OR data covering lockdown only partially |
| 5. Is there a chance of a reporting bias? | LR: data presented in its entirety (with respect to the research question of interest)<br>HR: data presented selectively |

### 6. Risk-of-bias assessment results

S1 Fig**. 1** shows the risk-of-bias assessment of surveys related to hepatitis, tuberculosis and malaria. Surveys related to HIV are assessed in S1 Fig. 2. RoB assessment of other studies concerning hepatitis and tuberculosis is shown in S1 Fig**. 3**, and concerning malaria and HIV in S1 Fig**. 4**. As to modelling studies, the main reason for concerns was the missing uncertainty analysis (for HIV [35-38], for malaria [35, 37, 39], for tuberculosis [35, 40], for viral hepatitis [41]). Two studies did not used a dynamic model ([37] for HIV, [41] for viral hepatitis). One preprint for HIV [42] did not contain a sufficient description of the model and underlying data sources, but the results seemed to be extended and later published in [43]. Similarly, the description of the data sources behind the model for tuberculosis used in [35] did not clearly state to what extent real world data and official records were utilized.

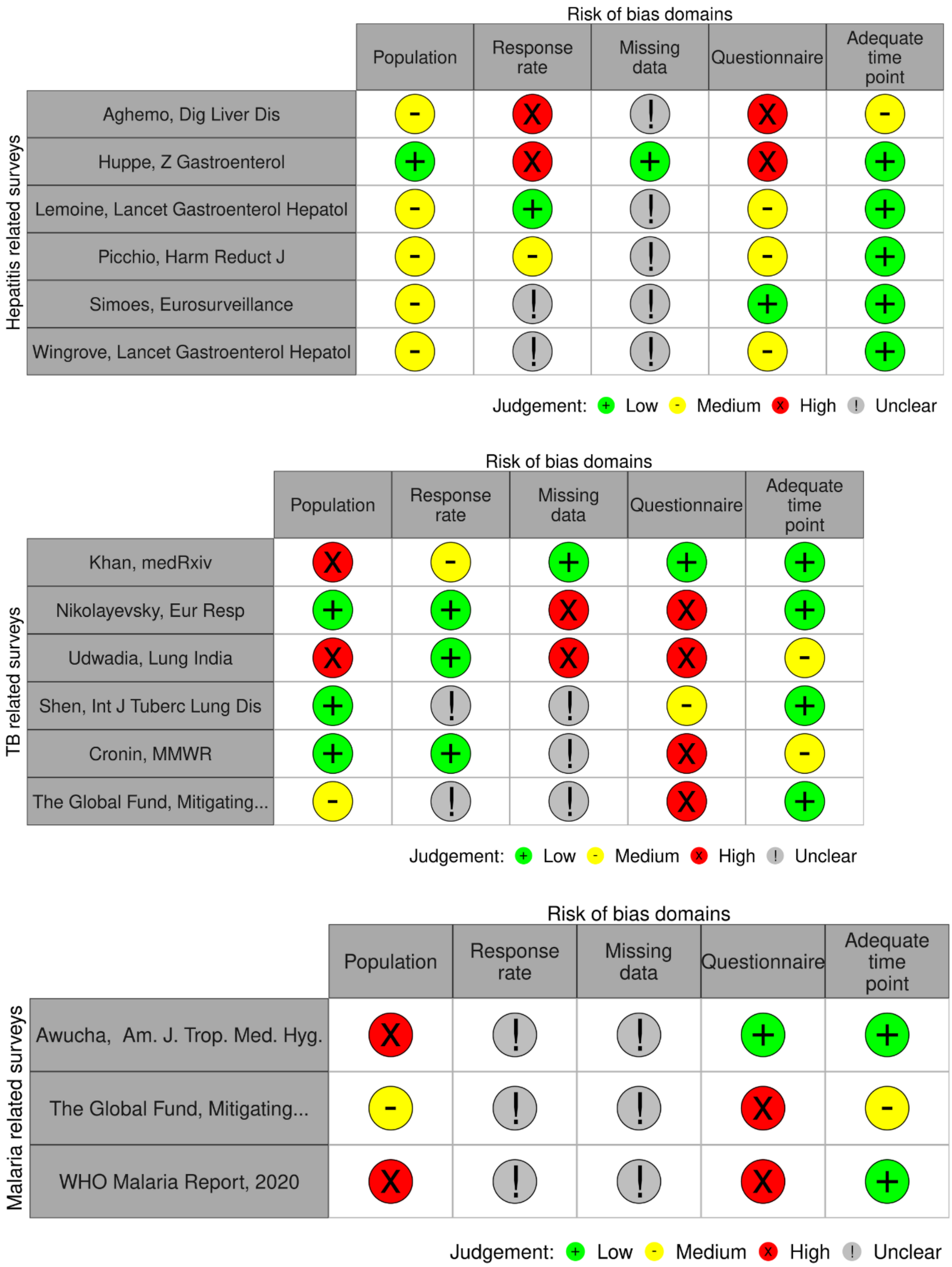

**S1 Fig. 1 Risk-of-bias assessment for surveys related to viral hepatitis, tuberculosis and malaria.**

| HIV related surveys | Risk of bias domains: Population | Response rate | Missing data | Questionnaire | Adequate time point |
|---|---|---|---|---|---|
| Bogart, J Acquir Immune Defic Syndr | - | - | X | - | + |
| Brawley, AIDS 2020 | X | X | ! | X | ! |
| Chow, Open forum infect dis | - | X | ! | X | + |
| Dawson, KFF report | + | X | ! | + | + |
| Dyer, AIDS and Behaviour | - | X | + | - | - |
| Guo, Zhonghua Liu Xing Bing Xue Za Zhi | X | X | ! | - | - |
| Hammoud, J Acquir Immune Defic Syndr | - | - | + | X | - |
| Hochstatter, AIDS and behaviour | - | X | ! | - | - |
| Kalichman, J Acquir Immune Defic Syndr | X | ! | + | + | X |
| Khan, medRxiv | X | ! | + | + | + |
| Kowalska, IJID | - | + | ! | X | - |
| Lamontagne, AIDS 2020 | X | ! | ! | + | + |
| Linnemayr, AIDS and Behaviour | - | - | + | X | - |
| Lourida, HIV Glasgow | X | ! | ! | X | + |
| Pampati, medRxiv | - | X | - | X | + |
| Picchio, Harm Reduct J | - | - | ! | - | + |
| Qiao, AIDS Care | X | X | ! | X | + |
| Rao, medRxiv | X | X | - | - | + |
| Reyniers, Sex Transm Infect | X | X | X | + | - |
| Sanchez, AIDS and Behaviour | X | X | ! | + | - |
| Santos, AIDS and Behaviour | X | X | ! | - | + |
| Siewe Fodjo, J Acquir Immune Defic Syndr | X | X | + | + | + |
| Simões, Euro Surveill | - | ! | ! | + | + |
| Stephenson, AIDS and behaviour | X | X | + | + | + |
| Sun, J Int AIDS Soc | X | X | + | - | - |
| The Global Fund reports | - | ! | ! | X | + |
| Torres, AIDS and Behaviour | X | X | + | X | + |
| WHO Pulse Survey | - | ! | + | + | + |

Judgement: + Low - Medium X High ! Unclear

**S1 Fig. 2 Risk-of-bias assessment of surveys related to HIV.**

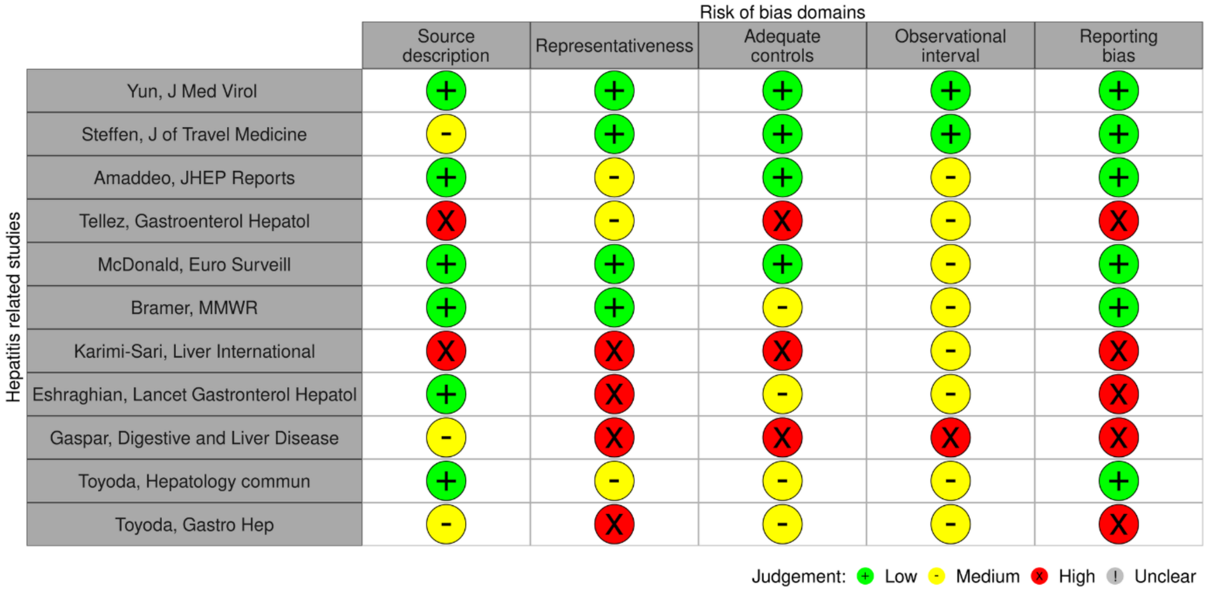

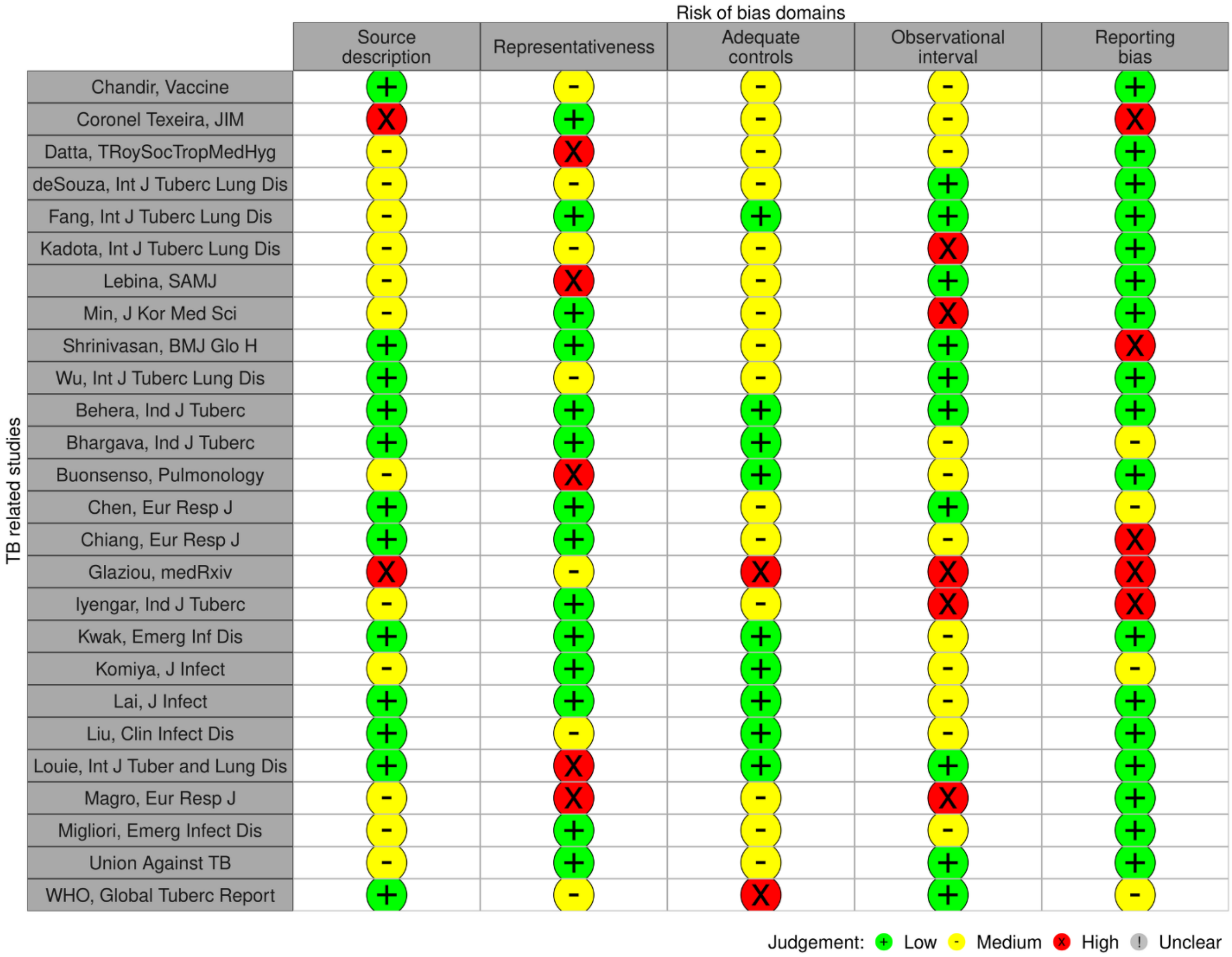

**S1 Fig. 3 Risk-of-bias assessment of quantitative studies related to viral hepatitis and tuberculosis.** Note that representativeness is assessed with respect to the global picture and is therefore high if only single centre results are reported.

Risk of bias domains

Malaria related studies

| Study | Source description | Representativeness | Adequate controls | Observational interval | Reporting bias |
|---|---|---|---|---|---|
| Bell, The Am J Trop Med Hyg | + | + | - | X | + |
| Buonsenso, medRxiv | + | X | + | - | + |
| Buonsenso,Frontiers in Pediatrics | + | X | - | - | + |
| Penjor, Malaria J | X | + | - | + | + |
| Ranaweera, Malaria J | X | + | - | + | X |
| Thapa,Research Square | + | + | - | + | + |
| Guerra, BMJ Global Health | X | X | - | + | X |
| Feldman,Research Square | X | X | X | + | X |
| RBM Partnership to End Malaria | X | + | X | + | X |
| Torres, Am J Trop Med Hyg | - | - | + | + | + |
| WHO Malaria Report 2020 | X | - | + | + | X |

Judgement: + Low - Medium X High ! Unclear

Risk of bias domains

HIV related studies

| Study | Source description | Representativeness | Adequate controls | Observational interval | Reporting bias |
|---|---|---|---|---|---|
| Bogdanic, HIV Glasgow | + | + | + | + | X |
| Chia, Sex Transm Infect | + | + | + | + | + |
| Chow, J Acquir Immune Defic Syndr | + | X | + | + | + |
| Chow, Lancet HIV | - | X | + | + | + |
| Darcis, Epid anf Infection | + | X | + | + | + |
| Davey, Lancet HIV | - | X | + | + | + |
| Ejima, medRxiv | + | + | + | + | + |
| Giuliani, Sex Transm Infect | X | X | X | + | + |
| Hickey, AIDS | + | X | + | + | + |
| Jensen, S Afr Med J | + | + | + | + | + |
| Junejo, Lancet HIV | - | X | + | + | + |
| Krakower, AIDS 2020 | + | X | - | - | + |
| Lagat, AIDS and Behaviour | - | - | + | + | + |
| Maatouk, Sex Transm Infect | X | X | - | + | X |
| Madhi, S Afr Med J | - | + | + | + | + |
| Mitsuya, Glob Health Med | + | + | + | + | + |
| Odinga, Gates Open Res | - | - | + | + | + |
| Ogirima, AIDS 2020 | X | X | | + | + |
| Panagopoulos, HIV Glasgow | - | X | - | + | + |
| Qiao, AIDS and Behaviour | + | - | | X | + |
| Quiros-Roldan, AIDS Res Ther | + | X | + | + | + |
| Sánchez-Rubio, Sex Transm Infect | X | - | + | + | X |
| Shi, Research Square | + | - | + | + | + |
| Siedner, BMJ Open | + | - | + | + | + |
| Spinelli, AIDS | - | X | + | + | + |
| Stanford, AIDS and Behaviour | + | X | X | + | + |
| Traeger, AIDS | + | + | + | + | + |
| Tun, AIDS 2020 | X | X | - | + | + |
| UNAIDS reports | X | + | X | + | X |
| Union Against TB | + | + | - | + | + |

Judgement: + Low - Medium X High ! Unclear

**S1 Fig. 4 Risk-of-bias assessment of quantitative studies related to malaria and HIV.** Note that the empty grey circles in case of HIV mean “not applicable”. Note that representativeness is assessed with respect to the global picture and is therefore high if only single centre results are reported.

### 7. Tuberculosis

Here we show the detailed findings quantifying the indirect impact of COVID-19 on TB. **S1 Fig. 5** shows how the numbers of notifications changed in Americas and Europe. An overview of all the identified studies with respect to the outcome and the type of evidence or of data sources is shown in **S1 Table 6**. Further tables detail the impact of COVID-19 on TB notifications(**S1 Table 7**), TB-related clinical outcomes(**S1 Table 8**), and TB-related service provision(**S1 Table 9**), as well as the projected impact on excess cases and deaths from the modelling studies(**S1 Table 10**). The entries in the tables are organized according to their geographical location, so a study may appear several times within a table, if it reports on several locations.

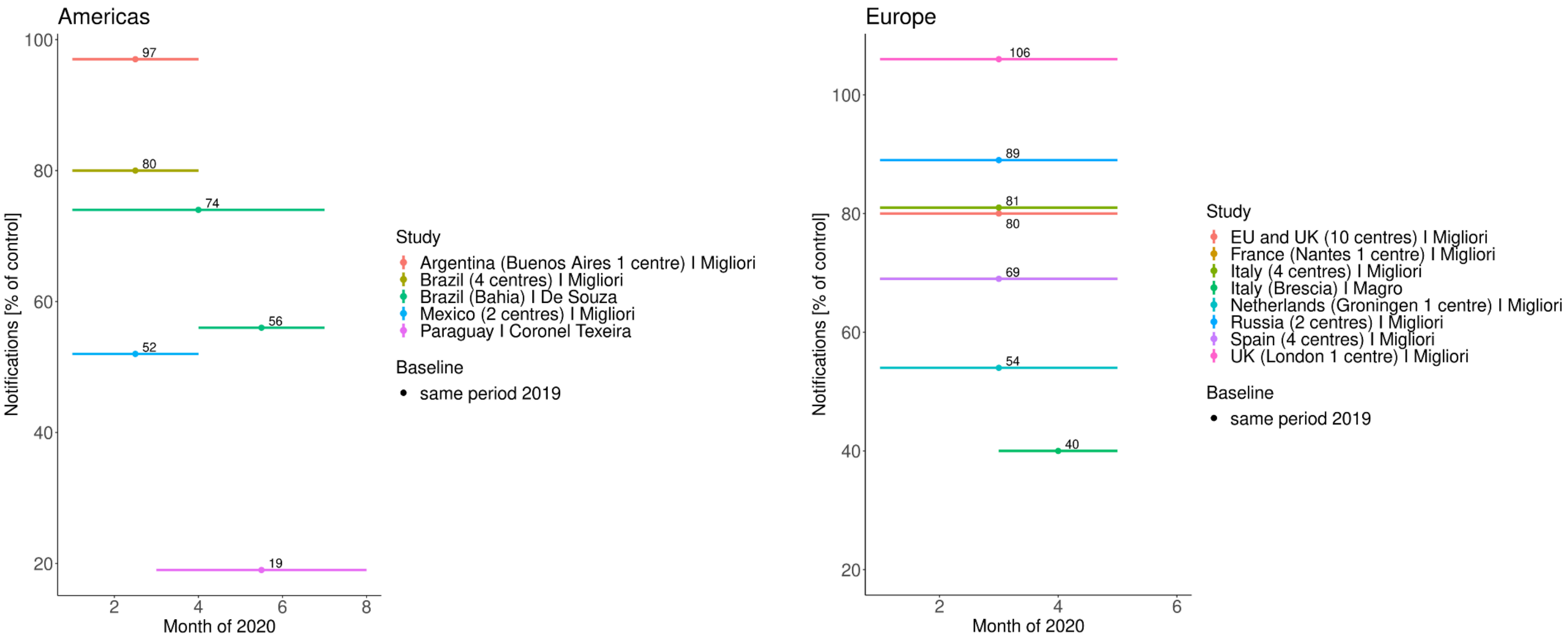

**S1 Fig. 5 TB notifications in Americas and Europe.** Overview of notifications registered over a certain period (depicted by the horizontal lines) as percentage of notifications in a control period (referred to as Baseline in the legend). Thus, a value of 20% means a relative decrease of 80%.

**S1 Table 6 Overview of studies related to TB with respect to the outcome and type of evidence/data source.**

| | National/ regional data | Hospital/program records | Surveys | Grey literature |
|---|---|---|---|---|
| **Impact on services and service provision** | Louie[44], Chandir[45], Shrinivasan[46], Wu[47] | Migliori[48] | Cronin[49], Khan[50], Nikolayevsky[51], Shen[52], Udwadia[53] | Global Fund[54], WHO[55] |
| **Notifications/cases** | Behera[56], Bhargava[57], Chen[58], Chiang[59], Coronel Teixera[60], Fang[61], Glaziou[40], Iyengar[62], Kwak[63], Komiya[64], Lai[65], Liu[66], Mohammed[67], de Souza[68], Wu[47] | Buonsenso[69], Datta[70], Magro[71], Migliori[48], Kadota[72], Lebina[73], Min[74] | Shen[52] | Union[75], WHO[55] |
| **Clinical outcomes** | Liu[66], Mohammed[67], Shrinivasan[46], Wu[47] | Magro[52], Min[74] | | Union[75] |
| **Modeling studies** | Bhargava[57], Cilloni[76], Glaziou[40], Kadota[72], McQuaid[77], Hogan[35] | | | STOP TB Partnership[78] |

**S1 Table 7 Overview of findings describing the impact of COVID-19 on TB notifications.** NNV = no numerical value.

| Author | Location | Data source | COVID time interval | Covid data | Control interval 1 | Control data 1 | Control interval 2 | Control data 2 | Other control intervals | Other control data | Data type | Impact/ summary |
|---|---|---|---|---|---|---|---|---|---|---|---|---|
| **ASIA** | **ASIA** | | | | | | | | | | | |
| Behera[56] | India | national | 20/3-3/7/20 | 362604 | 20/3-3/7/19 | 753087 | 20/3-3/7/18 | 679222 | 20/3-3/7/17 | 56216 | Notifications | -51.9% |
| | India | national | 22/3-12/4/20 | 11367 | 2-22/3/20 | 45875 | | | | | average weekly | -75.2% |
| Bhargava[57] | India | national | 25/3-19/5/20 | 154258 | 29/1-24/3/20 | 380205 | | | | | Notifications | -59% |
| Iyengar[62] | India | national | 4/20 | NNV | 4/19 | NNV | | | | | Notifications | -78% |
| Datta[70] | Pataudi block, Gurgaon, India | Program data | 3/20 | 46* | 3/19 | 51* | | | | | Cases | -9.8% |
| | | Program data | 4/20 | 40* | 4/19 | 62* | | | | | Cases | -35.5% |
| | | Program data | 5/20 | 19* | 5/19 | 85* | | | | | Cases | -77.6% |
| Chen[58] | China | National | 1/20 | 67882 | 1/19 | NNV | | | | | Notifications | -24% |
| | | National | 2/20 | 44933 | 2/19 | NNV | | | | | Notifications | -39% |
| | | National | 3/20 | 73427 | 3/19 | NNV | | | | | Notifications | -25% |
| | | National | 4/20 | 85684 | 4/19 | NNV | | | | | Notifications | -15% |
| | | National | 5/20 | 83385 | 5/19 | NNV | | | | | Notifications | -13% |
| Shen[52] | China | Hospital (13) | 1-3/20 | NNV | 1-3/19 | NNV | | | | | Notifications | -25% |
| | China | | 4/20 | NNV | 4/19 | NNV | | | | | Notifications | -15% |

| | | | | | | | | | | | | |
|---|---|---|---|---|---|---|---|---|---|---|---|---|
| Wu[47] | Shanghai, China | National | 24/1-26/3/20 | 2.4 (2.2-2.6) | 1/11/19 - 23/1/20 | 4.6 (4.3-4.8) | | | | | incidence per 1.000.000 person weeks | -47.8% |
| | | National | 24/1-26/3/20 | 2.4 (2.2-2.6) | 27/3 - 28/5/20 | 4.5 | | | | | incidence per 1.000.000 person weeks | -46.7% |
| Chiang[59] | China | National | 2/20 | NNV | 2/19 | NNV | | | | | | >-20% |
| | Malaysia | National | 1-5/20 | NNV | 1-5/19 | NNV | | | | | | -9% |
| Glaziou[40] | China | Not given | 2/20 | NNV | 2/20 | NNV | | | | | | -20% |
| | India | National | 2-22/3/20 | 45875 | 23/3-12/4/20 | 11367 | | | | | average weekly | -75% |
| | Indonesia | Not given | 3/20 | NNV | 1/20 | NNV | | | | | | -68% |
| Komiya[64] | Japan | National | 1/20 | 950* | 1/19 | 1000* | 1/18 | 1010* | 1/17 | 1100* | Notifications | -5% |
| | | | 2/20 | 900* | 2/19 | 1060* | 2/18 | 1065* | 2/17 | 1210* | Notifications | -15.1% |
| | | | 3/20 | 980* | 3/19 | 1100* | 3/18 | 1260* | 3/17 | 1265* | Notifications | -10.9% |
| | | | 4/20 | 830* | 4/19 | 1150* | 4/18 | 1135* | 4/17 | 1190* | Notifications | -17.8% |
| Kwak[63] | South Korea | National | weeks 1-8/20 | 498 | weeks 1-8/2015-19 | 594 | | | | | average weekly | -16% |
| | | National | weeks 9-18/20 | 390 | weeks 9-18/2015-19 | 655 | | | | | average weekly | -40% |
| Min[74] | South Korea | PPM data | 4-6/20 | 4893 | 7-9/19 | 6066 | | | | | notifications | -19% |
| Fang[61] | Taiwan | National | 1-8/20 | 5446 | 1-8/19 | 5907 | 1-8/2018 | 6280 | 1-8/2017 | 6623 | notifications | -7.8% |
| Lai[65] | Taiwan | National | weeks 1-20/20 | 2662 | Weeks 1-20/19 | 3307 | weeks 1-20/18 | 3512 | weeks 1-20/17 | 3563 | notifications | -19% |
| Liu[66] | Jiangsu, China | Regional | 1-5/20 | 6749 | 1-5/19 | 10620 | 1-5/15 | 14180 | | | notifications | -36% |
| Migliori[48] | Sidney, Australia | Hospital (1) | 1-4/20 | 52 | 1-4/19 | 35 | | | | | Cases | 48.57% |
| | Singapore | Hospital (1) | 1-4/20 | 16 | 1-4/19 | 31 | | | | | Cases | -48.39% |
| **AFRICA** | | | | | | | | | | | | |
| Buonsenso[69] | Western Area, Sierra Leone | Hospital (1) | 4/2020 | 5 | 4/19 | 15 | 4/2018 | 13 | | | Cases | -67% |
| | | | 4/2020 | 9 | 4/19 | 32 | 4/2018 | 30 | | | Suspected cases | -72% |
| Union[75] | Nairobi, Kenya | Hospital (18) | 3-8/2020 | 1334 | 3-8/19 | 2041 | | | | | Cases | -34.6% |
| | Lilongwe, Malawi | Hospital (8) | 3-8/2020 | 745 | 3-8/19 | 962 | | | | | Cases | -22.6% |
| | Harare, Zimbabwe | Hospital (10) | 3-8/2020 | 376 | 3-8/19 | 504 | | | | | Cases | -25.4% |
| | Nairobi, Lilongwe, Harare | | 3-8/2020 | 2455 | 3-8/19 | 3507 | | | | | Cases | -30% |
| | Nairobi, Kenya | Hospital (18) | 3-8/2020 | 8195 | 3-8/19 | 17500 | | | | | Suspected cases | -53.2% |

| | | | | | | | | | | | | |
|---|---|---|---|---|---|---|---|---|---|---|---|---|
| | Lilongwe, Malawi | Hospital (8) | 3-8/2020 | 3270 | 3-8/19 | 6496 | | | | | Suspected cases | -49.7% |
| | Harare, Zimbabwe | Hospital (10) | 3-8/2020 | 1057 | 3-8/19 | 1474 | | | | | Suspected cases | -28.3% |
| | Nairobi, Lilongwe, Harare | | 3-8/2020 | 12522 | 3-8/19 | 25470 | | | | | Suspected cases | -50.8% |
| Migliori[48] | Nairobi, Kenya | Hospital (1) | 1-4/2020 | 3733 | 1-4/19 | 4270 | | | | | Cases | -12.6% |
| | Zinder, Niger | Hospital (1) | 1-4/2020 | 594 | 1-4/19 | 704 | | | | | Cases | -15.6% |
| | Sierra Leone | Hospital (3) | 1-4/2020 | 114 | 1-4/19 | 135 | | | | | Cases | -16.7% |
| Mohammed[67] | Dere Dawa, Ethiopia | | 4-6/2020 | 110 | 1-3/20 | 270 | 10-12/2019 | 342 | 7-9/2019 | 326 | Notifications | -59% |
| Kadota[72] | Uganda | Hospital (58) | 30/3-2/5/20 | 517 (5 weeks) | 13/1-22/3/20 | 2045 (10 weeks) | | | | | Notifications | -43% |
| | | | 30/3-2/5/20 | 95-115 | 13/1-22/3/20 | 187-236 | | | | | Notifications | -43% |
| Lebina[73] | Johannesburg, South Africa | Hospital (1) | 27/3-30/9/20 | 3 (2-4) | 1/1/18-26/3/20 | 14 (5-25) | | | | | Average monthly cases | -78.6% |
| | | | 27/3-30/9/20 | 10 in 1000 | 1/1/18-26/3/20 | 18 in 1000 | | | | | TB cases per paediatric admission | |
| **AMERICAS** | | | | | | | | | | | | |
| Coronel Texeira[60] | Paraguay | National | 03-08/20 | NNV | 03-08/19 | NNV | | | | | Notifications | -80.8% |
| De Souza[68] | Bahia, Brazil | | 1-7/20 | 2094 | 1-7/19 | 2844 | | | | | Notifications | -26.4% |
| | | | 4-7/20 | 938 | 4-7/19 | 1663 | | | | | Notifications | -43.5% |
| Migliori[48] | Mexico | Hospital (2) | 1-4/20 | 83 | 1-4/19 | 158 | | | | | Cases | -47.5% |
| | Buenos Aires, Argentina | Hospital (1) | 1-4/20 | 76 | 1-4/19 | 78 | | | | | Cases | -2.6% |
| | Brazil | Hospital (4) | 1-4/20 | 268 | 1-4/19 | 337 | | | | | Cases | -20.5% |
| **EUROPE** | | | | | | | | | | | | |
| Magro[71] | Brescia, Italy | | 1/3-30/4/20 | 6 | 1/3-30/4/19 | 15 | | | | | Cases | -60% |
| Migliori[48] | Russia | Hospital (2) | 1-4/20 | 941 | 1-4/2019 | 1059 | | | | | Cases | -11% |
| | EU and UK [Italy (4), Spain (3), France (1), UK (1), the Netherlands (1)] | Hospital (10) | 1-4/20 | 399 | 1-4/2019 | 498 | | | | | Cases | -20% |

| | | | | | | | | | | | | |
|---|---|---|---|---|---|---|---|---|---|---|---|---|
| | Italy | Hospital (4) | 1-4/20 | 215 | 1-4/19 | 266 | | | | | Cases | -19.2% |
| | Spain | Hospital (3) | 1-4/20 | 55 | 1-4/19 | 80 | | | | | Cases | -31.2% |
| | Nantes, France | Hospital (1) | 1-4/20 | 24 | 1-4/19 | 35 | | | | | Cases | -31.4% |
| | Groningen, Netherlands | Hospital (1) | 1-4/20 | 20 | 1-4/19 | 37 | | | | | Cases | -46% |
| | London, UK | Hospital (1) | 1-4/20 | 85 | 1-4/19 | 80 | | | | | Cases | +6.3% |

*Values are approximated from graph via WebPlotDigitizer (https://apps.automeris.io/wpd/)

**S1 Table 8 Overview of findings regarding indirect impact of COVID-19 on different clinical outcomes related to TB.** DOTS= directly-observed therapy schedules, LTFU= loss to follow up.

| Author | Location | Data item | COVID time interval | COVID data | Control time intervals | Control data |
|---|---|---|---|---|---|---|
| **ASIA** | | | | | | |
| Liu[66] | Jiangsu, China | treatment completion | 1-5/2020 | 53.2% | 1-5/2019 | 61.2% |
| | | treatment success | 5/2020 | 89% | 2015-2020 | >90% |
| | | MDR screening rate - high risk | 5/2020 | 84% | 5/2019 | 99% |
| | | MDR screening rate - new diagnoses | 5/2020 | 81% | 5/2019 | 98% |
| Wu[47] | Shanghai, China | Sputum smears | 24/1-26/3/2020 | 86.2% | 1/11/2019 - 23/1/2020 | 95.7% |
| | | Xpert MTB/RIF | 24/1-26/3/2020 | 66.9% | 1/11/2019 - 23/1/2020 | 79.5% |
| | | diagnostic delay | 24/1-26/3/2020 | 18.5 (IQR 9-33) | 1/11/2019 - 23/1/2020 | 15 (IQR 6-29) |
| Min[74] | South Korea | Sputum smear test coverage | 4-6/2020 | 94.4% | 7-9/2019 | 94.6% |
| | | Sputum culture coverage | 4-6/2020 | 94.1% | 7-9/2019 | 94.2% |
| | | Treatment adherence | 4-6/2020 | 93.3% | 7-9/2019 | 93.4% |
| | | Drug-susceptibility test coverage | 4-6/2020 | 93.6% | 7-9/2019 | 93.6% |
| | | LTFU | 4-6/2020 | 2.1% | 7-9/2019 | 2.6% |
| | | Treatment success | 4-6/2020 | 84.1% | 7-9/2019 | 90.6% |
| Shrinivasan[46] | India | BCG vaccination | 3/2020 | NNV | 1/2020 | NNV |
| | | DOTS patients | 4/2020 | -50% | 2/2020 | 1 |
| | | Treatment success | 6/2020 | NNV | 6/2019 | NNV |
| | | TB hospitalisations | 6/2020 | -66% | 6/2019 | 1 |
| Chandir[45] | Sindh, Pakistan | BCG - fixed location | 23/03-09/05/2020 | -40.6% | 23/09/2019 - 22/03/2020 | 1 |
| | | BCG – outreach | 23/03-09/05/2020 | -83.5% | 23/09/2019 - 22/03/2020 | 1 |
| | | BCG - total | 23/03-09/05/2020 | -56.6% | 23/09/2019 - 22/03/2020 | 1 |
| **AFRICA** | | | | | | |
| Union[75] | Kenya, Nairobi | treatment success | 3-8/2020 | 64% | 3-8/2019 | 67.8% |
| | Lilongwe, Malawi | treatment success | 3-8/2020 | 96.5% | 3-8/2019 | 96.3% |
| | Harare, Zimbabwe | treatment success | 3-8/2020 | 63.3% | 3-8/2019 | 80.1% |
| | Nairobi, Lilongwe, Harare | treatment success | 3-8/2020 | 72.3% | 3-8/2019 | 77.7% |
| Mohammed[67] | Dere Dawa, Ethiopia | Cure | 4-6/2020 | 11 | 1-3/20; 10-12/19; 7-9/19 | 75; 79; 85 |
| | | treatment completion | 4-6/2020 | 67 | 1-3/20; 10-12/19; 7-9/19 | 92; 147; 194 |

| | | | | | | |
|---|---|---|---|---|---|---|
| | | LTFU | 4-6/2020 | 5 | 1-3/20; 10-12/19; 7-9/19 | 3; 4; 2 |
| | | Death | 4-6/2020 | 6 | 1-3/20; 10-12/19; 7-9/19 | 10; 5; 10 |
| | | Treatment failure | 4-6/2020 | 0 | 1-3/20; 10-12/19; 7-9/19 | 0; 2; 0 |
| **EUROPE** | | | | | | |
| Magro[71] | Brescia, Italy | LTFU | 1/3-30/4/2020 | 7/65 | 1/3-30/4/2019 | 2/76 |
| | | Deaths | 1/3-30/4/2020 | 3/65 | 1/3-30/4/2019 | 0/76 |
| | | Completed treatment | 1/3-30/4/2020 | 10/65 | 1/3-30/4/2019 | 11/76 |
| | | patients newly entering TB care | 1/3-30/4/2020 | 65 | 1/3-30/4/2019 | 76 |

**S1 Table 9 Overview of published evidence regarding impact of COVID-19 on TB-related service provision.** IP= in-patient, OP=out-patient, PPE = personal protective equipment.

| **Author** | **Location** | **Data item** | **Time interval (control interval)** | **COVID data (control data)** | **Control interval** | **Control data** |
|---|---|---|---|---|---|---|
| WHO[55] | 184 countries | fewer health facilities providing OP care for tb | 1-6/2020 | 32/184, 7/30 high burden | | |
| | | fewer health facilities providing OP care for drug-resistant tb | 1-6/2020 | 21/184, 4/30 high burden | | |
| | | fewer hospitals providing IP care for tb | 1-6/2020 | 35/184, 9/30 high burden | | |
| | | fewer hospitals providing IP care for drug-resistant tb | 1-6/2020 | 33/184, 9/30 high burden | | |
| | | reduced number of OP visits for patients with tb | 1-6/2020 | 127/184, 28/30 high burden | | |
| | | reallocation of staff | 1-6/2020 | 85/184, 20/30 high burden | | |
| | | reallocation of funding | 1-6/2020 | 52/184, 14/30 high burden | | |
| | | reallocation of GeneXpert machines | 1-6/2020 | 43/184, 13/30 high burden | | |
| | | people with Tb asked to self-isolate at home | 1-6/2020 | 93/184, 14/30 high burden | | |
| Global Fund[54] | 104 country agents | Tb service disruption | 01/06/2020 | 78%, 17% high/ very high | | |
| | | lab services disruption | 01/06/2020 | 20% high or very high | | |
| | | shortage of medical supplies and treatment | 01/06/2020 | 9% | | |
| Khan[50] | 64 LMICs | Access for patients to healthcare facility | 12/5 - 5/8/2020 | 40% very hard/impossible | | |
| | | Access for provider to healthcare facility | 12/5 - 5/8/2020 | 37% very hard/impossible | | |
| | | Access for patients to non-medical support | 12/5 - 5/8/2020 | 31% very hard/impossible | | |
| | | TB medical treatment provision | 12/5 - 5/8/2020 | 11% very hard/impossible | | |
| | | Diagnostic service provision | 12/5 - 5/8/2020 | 29% very hard/impossible | | |
| Cronin[49] | USA | staffing capacity | 4/2020 | 60-72% partial/ high impact | | |
| | | essential services for tb patients | 4/2020 | 52% partial/ high impact | | |
| | | essential services for latent tb patients | 4/2020 | 68% partial/ high impact | | |
| | | contact tracing | 4/2020 | 64% partial/ high impact | | |
| | | testing/treatment of latent tb in populations at risk | 4/2020 | 74% partial/ high impact | | |
| | | case reporting and surveillance | 4/2020 | 58% partial/ high impact | | |
| | | program evaluation | 4/2020 | 74% partial/ high impact | | |
| | | education and training | 4/2020 | 94% partial/ high impact | | |
| | | challenges obtaining medication | 4/2020 | 15% | 3/2020 | 7% |
| | | reallocation of resources (e.g. PPE) | 4/2020 | 12% | 3/2020 | 7% |

| | | | | | | |
|---|---|---|---|---|---|---|
| Louie[44] | San Franscisco, USA | B-notification immigrant evaluations | 2-5/2020 | 19 | 2-5/19; 2-5/18 | 76; 84 |
| Shen[52] | China | change in no. of beds for TB | 1-3/2020 | -75.7% | 1-3/2019 | 1 |
| | | change in no. of beds for TB | 4/2020 | -43.6% | 4/2019 | 1 |
| | | change in no. of staff for TB | 1-3/2020 | -28.5% | 1-3/2019 | 1 |
| | | change in no. of staff for TB | 4/2020 | -10% | 4/2019 | 1 |
| | | hospitals setting strict indications for hospital admission | 1-3/2020 | 38.5% | 1-3/2019 | 0 |
| | | hospitals setting strict indications for hospital admission | 4/2020 | 23.1% | 4/2019 | 0 |
| | | hospitals setting restrictions for maximum OP visits | 1-3/2020 | 30.8% | 1-3/2019 | 23.1% |
| | | hospitals setting restrictions for maximum OP visits | 4/2020 | 23.1% | 4/2019 | 23.1% |
| | | change in no. of OP visits | 1-3/2020 | -34% | 1-3/2019 | 1 |
| | | change in no. of OP visits | 4/2020 | -20% | 4/2019 | 1 |
| | | change in no. of admissions | 1-3/2020 | -30% | 1-3/2019 | 1 |
| | | change in no. of admissions | 4/2020 | -18% | 4/2019 | 1 |
| Wu[47] | Shanghai, China | TB clinics closed | 24/1-26/3/2020 | 6/26 | | |
| | | Tertiary centres halving hospital beds | 24/1-26/3/2020 | 2/4 | | |
| Udwadia[53] | Mumbai, India | problems collecting drugs | 24/3/2020- | 78% | | |
| | | problems finding lab for ECG/bloods | 24/3/2020- | 64% | | |
| | | difficulties with transport | 24/3/2020- | 21% | | |
| | | issues with administration of injectables | 24/3/2020- | 7% | | |
| Nikolayevsky[51] | EU reference laboratories | Impact on services significant/ very significant | 11/3-11/4/2020 | 17/30 | | |
| | | Impact on services significant/ very significant | 11/4-11/5/2020 | 14/30 | | |
| | | Impact on services significant/ very significant | 11/5-11/6/2020 | 10/30 | | |
| | | Impact on training significant/ very significant | 11/3-11/4/2020 | 23/30 | | |
| | | Impact on research and development significant/ very significant | 11/3-11/4/2020 | 15/30 | | |
| | | >25% reduction in workload in primary diagnostic services | 11/3-11/4/2020 | 16/27 | | |
| | | >25% reduction in workload in reference services | 11/3-11/4/2020 | 12/29 | | |

**S1 Table 10 Overview of the modelling studies regarding indirect impact of COVID-19 on TB.**

| Author | Location | Scenario | Excess cases over 5 years | Excess deaths over 5 years |
|---|---|---|---|---|
| Cilloni[76] | Worldwide | 3m lockdown, 10m restoration; TB transmission -50% | +8% (4.702.300) | + 12.3% (1044800) |
| Cilloni[76]/ StopTB[78] | Worldwide | 3m lockdown, 10m restoration; TB transmission -10% | +10.7% (6.331.100) | +16% (1367300) |
| Glaziou[40] | Worldwide | Decrease in case detection by 25% over 3m | | +13% (1.660.000) |
| | Worldwide | Decrease in case detection by 50% over 3m | | +26% (3.320.000) |
| McQuaid[77] | China, India, South Africa | Low reduction in TB transmission, high demand on healthcare system | | +8-14% |
| Hogan[35] | One very high burden setting, one moderate burden setting | Pandemic suppression, high demand on health services | | +20% |

### 8. Viral hepatitis

Of 21 records eligible for inclusion and data extraction, 6 were surveys[27, 79-83], 3 modelling studies[41, 84, 85], 11 other studies [86-96] using mostly hospital or notifiable disease records, and one was a press article [97] based on hospital records. The details are presented in **S1 Table 11** (modelling studies), **S1 Table 12** (surveys) and **S1 Table 13** (other studies incl. grey literature sources). Note that the studies are grouped by outcomes and thus some may appear in the table several times, if they report on more than one outcome.

**S1 Table 11 Findings of the modelling studies related to indirect impact of COVID-19 on viral hepatitis.** RR= relative risk, HCC= hepatocellular carcinoma, HIC = high-income countries, UMIC = upper-middle-income countries, LMIC = lower-middle-income countries, LIC = low-income countries, UI = uncertainty interval.

| Author | Region | Baseline scenario | COVID-19 impacted scenario | Endpoint | Result | Comment |
|---|---|---|---|---|---|---|
| Lai et al. [41] | UK | 1-year mortality estimated based on data from 2012-2016 in England, scaled-up to the population of England aged 30+ (35 million individuals) | RR resulting from COVID-19 infection: 1.2, 1.5, 2<br>RR resulting from service change: 1.2, 1.5, 2<br>Infection rate over 1-year: 10%<br>Proportion affected by service change 30%, 70% | Excess deaths for incident liver cancers over a 1-year period | 55, 137, 274 at RR 1.2; 1.5; 2, respectively, applied to 10% infected and 30% affected by service change<br>110, 274, 548 at RR 1.2; 1.5; 2, respectively, applied to 10% infected and 70% affected by service change | Mixed direct and indirect effect of COVID-19.<br>Ad-hoc chosen RR representing the service interruption/change. |
| | | | | Excess deaths for prevalent liver cancers over a 1-year period | 26, 65, 131 at RR 1.2; 1.5; 2, respectively, applied to 10% infected and 30% affected by service change<br>52, 131, 262 at RR 1.2; 1.5; 2, respectively, applied to 10% infected and 70% affected by service change | |
| Abbas et al. [85] | 54 African countries | --- | 6 months of COVID-19, 6-month suspension of vaccination, no catch-up thereafter, unvaccinated children have the same risk of disease as children in a completely unvaccinated population | Benefit/risk ratio of HBV vaccination at 6, 10, 14 weeks (comparing excess COVID-19 deaths resulting from vaccination visits and deaths averted by vaccination up to 5 years of age) | Benefit/risk for household: 1 (95% UI: 0.1 -2)<br>Benefit/risk for vaccinated child: 686 (95% UI: 33 – 4688) | HBV vaccination is done together with vaccination against diphtheria, tetanus, pertussis, Haemophilus influenza type b, Streptococcus pneumonia, rotavirus. Considered together, the benefit/risk ratio for a household is 82 (14 – 261), supporting vaccination without doubt. |
| Blach et al. [84] | Global (110 countries) | Number of screened patients remains constant on the last known level after 2019, number of patients initiated on treatment annually drops to 50% over 5 years | No patients diagnosed or initiated on treatment in 2020, thereafter reverting to baseline scenario in 2020, resulting in 1-year offset | Excess prevalent infections 2020-2030 | 623 000 (95% UI: 609 000- 685 000) | No region projected to meet the target for incidence (90% reduction in new infections by 2030), only high-income countries projected to meet the target for mortality (65% reduction in liver-related deaths).<br>Excess liver related death only 50 600 by a 6-month delay scenario, and 25 300 by a 3-month delay scenario. |
| | | | | Excess incident HCV 2020-2030 | 121 000 (95% UI: 118 000 – 133 000) distributed between income groups as follows: HIC: 15%, UMIC: 28%, LMIC: 55%, LIC: 2% | |
| | | | | Excess incident HCC 2020-2030 | 44 800 (95% UI: 43 800 – 49 300) distributed between income groups as follows: HIC: 45%, UMIC: 23%, LMIC: 30%, LIC: 2% | |
| | | | | Excess liver related deaths 2020-2030 | 72 300 (95%UI: 70 600 – 79 400) distributed between income groups as follows: HIC: 41%, UMIC: 21%, LMIC: 35%, LIC: 3% | |

**S1 Table 12 Findings of the surveys related to indirect impact of COVID-19 on viral hepatitis.** Reported are changes in offer and uptake of hepatitis health services. DAA = direct-acting antiretroviral, LMIC= low-income and middle-income countries, HCC = hepatocellular carcinoma

| Author | Country | Population | Outcome | Baseline | COVID-19 period | N | Result | Comment |
|---|---|---|---|---|---|---|---|---|
| Hüppe et al[80] | Germany | Centres of the German hepatitis C registry | Liver consultations | ---- | Mar – May 2020 | 47 | 69% reduced or cancelled consultations<br>>50% appointments cancelled by pts | The offer of consultations went back to pre-covid state in July.<br>78% of the centres stated that the pandemic did not worsen the general care for pts with chronic HCV. |
| | | | Number of new pts | Mar – May 2019 | Mar – May 2020 | 64 | >6 new pts/month on average: 43.8% in 2019 vs 25% in 2020 | |
|  |  |  | Delayed diagnosis | ---- | Mar – May 2020 | 64 | HCC: 9.4%<br>Liver decompensation: > 22% |  |
| Simoes et al[79] | Countries of WHO European region | Providers of testing services reached through EuroTEST consortium network | Testing HBV | Mar – May 2019 | Mar – May 2020 | 47 | Severe decrease(>50% volume): 63% | Approximate percentages read off from the plots. |
| | | | | | Jun-Aug 2020 | 48 | Severe decrease(>50% volume): 15%<br>Decrease(>10%): 52%, Increase(>10%): 17% | |
| | | | Testing HCV | Mar – May 2019 | Mar – May 2020 | 66 | Severe decrease(>50% volume): 67% | |
| | | | | | Jun-Aug 2020 | 70 | Severe decrease(>50% volume): 19%<br>Decrease (>10%): 56%, Increase (>10%): 17% | |
| Aghemo et al[81] | Italy | Members of the Italian Assoc. for the Study of the Liver | Number of beds | Pre-covid | Apr 8 – May 3, 2020 Lockdown | 194 | Reduction reported by 33% | No quantitative assessment of the reductions, no pre-specified comparison interval.<br><br>26% respondents reported hepatology ward being converted to COVID-19 ward. |
| | | | Day hospital | | | | Reduction:47%<br>Complete interruption: 22% | |
| | | | Day service | | | | Reduction: 29%<br>Complete interruption: 24% | |
| | | | No change in outpatient activity | | | | Chronic hepatitis and compensated cirrhosis: 0.6%<br>HCC: 18%<br>Decompensated cirrhosis: 32% | |
| | | | HBV/HCV initiation of antiviral treatment | | | | Postponed: 23% | |
| | | | Cirrhosis screening | | | | Reduced: 46%, suspended: 20% | |
| | | | Liver transplant | | | | Pre-transplant assessment reduced: 30%<br>Post-transplant reviews postponed: 44% | |
| | | | HCC | | | | Reductions in patients starting treatment: 27%<br>No change in treatment activities: 34% | |
| Wingrove et al[27] | Global, 32 countries | Civil society organisations, other hepatitis service providers | Viral hepatitis testing | --- | Mar 30 – May 4, 2020 | 132 (48% USA) | 64% lack of access (reasons: closing of facilities: 46%, fear of Covid-19: 65%) | 123 of 131 (94%) reported their services had been effected by the crisis. |
| | | | Access to hepatitis medication | | | | Lack of access: LMIC: 52% (15/29) , USA 8% (5/64) | |
| | | | Reasons for the lack of access to hepatitis treatment | | | 64 | 50% (32/64): avoiding facilities due to COVID-19<br>41% (26/64): services redeployed to fight COVID-19<br>Outside USA: 55%(22/40): travel restrictions | |

| | | | | | | | | |
|---|---|---|---|---|---|---|---|---|
| Lemoine et al[83] | Burkina Faso, Tanzania, The Gambia | Retrospectively counted patients in main referral hospitals for chronic hepatitis HBV, HCV (3 from Burkina Faso, 1 from Tanzania, 1 from The Gambia) | Number of new cases | Jan 2020 | Apr 2020 | 5 | Burkina Faso: -71% (96 vs 28) , Tanzania: -95% (107 vs 5) , The Gambia: -83% (36 vs 6) | Rapid tests for HBV not disrupted, but shortage of nucleic acid tests for HBV DNA or HCV RNA.<br>Direct-acting antiretrovirals (DAAs) for HCV temporarily unavailable in Burkina Faso.<br><br>New patients and patients in follow-up in 2020 compared against those seen during the same period last year (Jan 1 to April 30, 2019) in The Gambia shown no significant fluctuation |
| | | | Number of follow-up patients | | | 5 | Burkina Faso: -73% (185 vs 50) , Tanzania: -77% (35 vs 8) , The Gambia: -98% (142 vs 15) | |
| | | | Outpatients visits | | | 5 | Reduction of more than 70%.<br>In Tanzania and The Gambia, clinics were temporarily closed at the end of March, 2020 in Burkina Faso clinics remained opened the whole time. | |
| Picchio et al[82] | Spain | Harm reduction service providers | Persons receiving DAA for HCV | Mar – Jun 2019 | Mar – Jun 2020 | 5 | Reduction of 82% (88 persons in 2019 vs 16 persons in 2020) | The average number of users decreased by 22% in 2020 (Jan-June) vs 2019 (from 292 to 215). |
| | | | HBV, HCV testing | | | 6 | Mostly decreases in testing rates per 1000 service users in March, April, May 200 compared to 2019, some centres reported great increase in June (year 2020 compared to 2019). | |

**S1 Table 13 Findings of studies relying on hospital or notifiable disease records, registries and databases, when assessing indirect impact of COVID-19 on viral hepatitis.**

| **Author** | **Country** | **Data source** | **Outcome** | **Baseline** | **COVID-19 period** | **Result** | **Comment** |
|---|---|---|---|---|---|---|---|
| **Vaccination** | | | | | | | |
| Bramer et al[90] | US Michigan State | Michigan Care Improvement Registry | HBV birth dose | May 2016, 2017, 2018, 2019 | May 2020 | No change | Less profound declines observed at ages 3, 7, 16, 19, 24 months.<br>Recommended vaccination includes HBV and HAV vaccines.<br>Size of cohorts: 9537 (in 2020), 9269 (average in 2016 – 2019) |
| | | | Completed recommended vaccination at 5 months of age | May 2016, 2017, 2018, 2019 | May 2020 | Declined from 66.6%, 67.4%, 67.3%, 67.9% during 2016-2019, respectively, to fewer than half 49.7% in May 2020 | |
| | | | Non-influenza vaccine doses administered | Jan – Apr 2018 and 2019 (averaged) | Jan – Apr 2020 | Children ≤18 years: decrease by 21.5%, children aged ≤24 months: decrease by 15.5% | |
| McDonald et al[92] | England | Electronic patient records:<br>The Phoenix Partnership (TPP) SystemOne | Delivered first doses of hexavalent vaccine (incl. HBV vaccine) to infants younger than 6 months | Week 1 – 17 in 2019 | Week 1-17 in 2020 | Week 1-17: -3.5% (-3.7% to -3.4%)<br>Week 1-9: -5.8% (-6.0% to -5.5%)<br>Week 10-12 (transition period): -4.4% (-4.8% to -4.0%)<br>Week 13-15 (physical distancing): -6.7% (-7.1% to -6.2%) | 67 116 in 2020, 69 568 in 2019<br><br>Modelling suggested a general decrease in vaccination in weeks 1 to 15 in 2020 compared with the same weeks in 2019, which did not |

| | | | | | | | |
|---|---|---|---|---|---|---|---|
| | | | | | | Week 16 -17: increase<br><br>(the 95% confidence intervals are calculated using the Wilson method)<br><br>Weeks 13-15: large regional variation, decrease of more than 10% up to increase of 17%. | accentuate on introduction of physical distancing, but reversed in week 15, with a percent increase in weeks 16 and 17 of 2020 compared to 2019.<br><br>No adjustment for eligible vaccine recipients; birth rates might play a role with less births. Coverage of vaccination is decreasing over previous years in general. |
| Yun et al[87] | South Korea | Korea Centers for Disease Control and Prevention, Press release April 22, 2020 | Vaccination coverage | Jan-Mar 2019 | Jan-Mar 2020 | Sustained:<br>HBV, 3 doses: 94.6% in 2019 vs 94.1% in 2020<br>HAV, 2 doses: 93.8% in 2019 vs 94.9% in 2020 | |
| Press article | India (unclear if country-wide or state) | Health Management Information System | Hepatitis B birth dose | Apr-May 2019 | Apr-May 2020 | Reduction of 19.4% in administered doses.<br><br>“19.4 per cent drop in Hepatitis-B birth doses administered and 31 per cent drop in vaccination sessions held in health facility and outreach sessions from April to June 2020 as compared to same period last year as per Health Management Information System (HMIS), in the light of the Covid-19 pandemic.” [citation from a website, see the cell to the right] | https://hepvoices.org/2020/09/19-4-drop-in-hepatitis-b-birth-doses-31-drop-in-vaccination-sessions-in-april-june/, accessed 30th Sept 2020 |
| **Incidence of HAV and HBV** | | | | | | | |
| Yun et al[87] | South Korea | National Notifiable Disease Surveillance System | Incidence of HAV | Jan – Jun 2015 to 2018 (averaged) | Jan – Jun 2020 | 1736 cases in 2020, corresponds to 14% decline in comparison to pre-years<br>Incidence rate difference significant during intensified social distancing measures (April, May 2020): -0.56 per 100 000 (95% CI: -0.80 - -0.32) | Poisson confidence interval.<br>Nationwide outbreak of HAV in 2019 is a reason for including only years 2015-2018 in the baseline. |
| | | | Incidence of HBV | Jan – Jun 2015 to 2019 (averaged) | Jan – Jun 2020 | 167 cases in 2020, no significant decline | |
| Steffen et al[88] | Switzerland | Federal Office of Public Health Bulletins | Incidence of HAV | Weeks 18-29 in 2016 to 2019 | Weeks 18-29 in 2020 | No significant decrease as compared to previous years<br>19 cases in 2020 correspond to 5% reduction in comparison to mean 2016-2019 (20 cases) | Switzerland in isolation from week 14 to week 24, incubation period of HAV: 4 weeks |

| | | | | | | | |
|---|---|---|---|---|---|---|---|
| **Number of patients visits** | | | | | | | |
| Toyoda et al[94] | Japan (Ogaki) | Electronic medical records | % of kept monthly scheduled visits for HCC surveillance | Jul 2019-Feb 2020 | Mar – May 2020 | Decline: 279 of 438(63.7%) visits kept in March-May 2020 vs 1236 of 1271 (97.2%) visits kept previously (p<0.0001).<br>Over 95% visits kept for each month up to February 2020, then decline: March: 79.7%, April: 63%, May: 49.1% | One centre, 935 patients with sustained virologic response, who did not have active HCC in July 2019 and continued visiting hospital for HCC surveillance.<br>Decline less pronounced in cirrhosis or advanced fibrosis patients and those with a history of HCC. Before February 2020 the majority of patients who cancelled did not reschedule; from March 2020, most patients did reschedule. |
| Toyoda et al.[95] | Japan (Ogaki), USA(Stanford), Singapore | Electronic medical records | Chronic HCV visits | Feb 1 – Mar 14, 2018 and 2019<br>Mar 15 – May 1, 2018 and 2019 | Feb 1 – Mar 14, 2020<br>Mar 15 – May 1, 2020 | Decreasing trend (p=0.022), separate analyses: Stanford: p=0.002, Ogaki: p=0.281 | One centre per country, all visits: 14 403, Stanford: 9345 , Ogaki: 3657 , Singapore: 1401<br>Significant decrease in total number of clinic visits from 2018 to 2019 to 2020 (p=0.038; from 5900 in 2018 to 5270 in 2019 to 3233 in 2020) in all three centres together; but: in separate analyses by site significant decrease only in Stanford (p-trend: 0.038) and Singapore (p-trend: 0.026) but not Ogaki.<br><br>p-values from a logistic regression, analysis not well described. |
| | | | Chronic HBV visits | | | Decreasing trend (p=0.003), separate analyses: Stanford: p=0.103, Ogaki: p=0.016, Singapore: p=0.001 | |
| | | | Visits of patients with HCC and/or cirrhosis | | | Decreasing trend not significant (p=0.110) | |
| | | | Liver imaging | | | Abdominal ultrasound: decreasing trend (p=0.003), separate analyses: Stanford: p=0.010, Ogaki: p=0.572, Singapore: p=0.753<br>CT/MRI: decreasing trend (p=0.007), separate analyses: Stanford: p=0.021, Ogaki: p=0.838, Singapore: p=0.012 | |
| **Liver related morbidities** | | | | | | | |
| Amaddeo et al[86] | France (Paris area) | Digital integrated care patient files | Number of patients with HCC presented to multidisciplinary tumor board | Mar 6 – Apr 17, 2019 | Mar 6 –Apr 17, 2020 | Decrease(221 in 2020 vs 304 in 2019)<br>Decrease over weeks (March 6 –March 26 vs March 27-April 17) in 2020 (p=0.034, Fisher's exact test). Less pronounced decrease for first diagnoses.<br>First diagnosis of HCC: larger tumor size (p=0.002, Mann-Whitney test) | Six academic hospitals in urban area of Paris, patients presented to multidisciplinary tumor board or with programmed surgical or radiological procedure with intention to treat in the studied periods: n=377 in 2019, n=293 in 2020.<br>Early lockdown. For 7% of patients in 2020 the treatment was not realized and so a reliable delay could not be calculated.<br>Most of the patients with a treatment delayed for more than 1 month have a recurrent HCC, suggesting perhaps that physicians |
| | | | Rate of treatment in patients with active HCC | | | Decrease: 56.7% (n =377) in 2019 vs. 43.7% (n = 293) in 2020, with a significant decrease during the second half of the time period in 2020 (p =0.018, Fisher's exact test). | |
| | | | Decision-to-treatment interval | | | No significant difference (2020 vs 2019).<br>Higher rate of patients with a delay longer than 1 month (21.5%, 63 cases in 2020 vs. 9.5%, 36 cases in 2019, respectively; p =0.001, chi-squared test). | |

| | | | | | | | |
|---|---|---|---|---|---|---|---|
| | | | | | | Recurrent HCC: higher rate of patients with delay above 1 month in 2020 (23.3%, 44 cases vs 4.7%, 11 cases, p<0.001, chi-squared test).<br>Delay or change in treatment strategy was associated only with the period 2020 in multivariate logistic regression (adjusted OR= 9.661 [95% CI: 2.85–32.72] ). Similar results for treatment delay above 1 month. | tended to delay treatment for patients, whose tumor biology they were familiar with and could adjust the delay accordingly. |
| Eshraghian et al[89] | Iran (Shiraz) | Patient charts | Hospital admissions for liver-related disorders | Feb 19 – Apr 30, 2019 | Feb 19 – Apr 30, 2020 | Decrease (230 in 2019 vs 124 in 2020), mean rate per day: 3.23(SD 1.33) in 2019 vs 5.32(SD 3.37) in 2020, IRR 1.85, 95% CI: 1.49-2.30 (Poisson regression) | One hepatobiliary referral centre,<br>IRR = incidence rate ratio<br>MELD = Model for End-Stage Liver Disease<br>Unclear what test some of the p-values are related to. |
| | | | Hospital stay duration | | | Mean duration longer during pandemic (5.32 days (SD 3.37) in 2019 vs 7.44 days (SD 5.42) in 2020, p<0.001) | |
| | | | MELD score for liver cirrhosis patients | | | Higher in 2020 (15.18 (SD 3.45) in 2019 vs 17.07 (SD 4.05) in 2020, p <0.001) | |
| Gaspar et al[91] | Portugal | Electronic health records and hospital discharge database | Hospital admissions for decompensated cirrhosis | Mar 2 – May 2, 2015 to 2019 | Mar 2 – May 2, 2020 | No difference (40 in 2020, median 38, range: 34-42 in previous years) | Tertiary academic hepatology centre<br>Unclear what tests the p-values are related to.<br>MELD = Model for End-Stage Liver Disease<br>CPD = Child-Purgh-Turcotte |
| | | | Length of hospital stay | | | No difference (6.5 days (range 1-27) in 2020 vs 8 days (range 1-58) previously, p=0.48) | |
| | | | In-hospital mortality | | | No difference (15% in 2020 vs 11.7% 2015-2019, p=0.57) | |
| | | | MELD score | | | No difference (14 (range 7-32) in 2020 vs 15 (range 7-32) previously) | |
| | | | CPD score | | | No difference (9(range 6-13) in 2020 vs 8 (range 5-14) previously) | |
| | | | Readmissions | | | More in 2020 (47.5% in 2020 vs 27.7% in 2019, p=0.014), associated with shorter index hospital stay. | |
| Tellez et al[96] | Spain | Organización Nacional de Transplantes. COVID-19: Impacto en la actividad de donación y trasplantes. 2020. | Liver transplantations | Jan, Feb 2020 | Mar, Apr, May 2020 | Decrease: 20-30 liver transplants weekly in Jan and Feb 2020 compared to less than 1/3 of that in March and April 2020 and back to 10-15 (half of initial level in 2020) in May 2020. | |
| **Research** | | | | | | | |
| Karimi-Sari et al[93] | Global | PubMed records | Number of records when searching for Hepatitis[tiab] | Jan-Mar 2019 | Jan-Mar 2020 | Decrease by 995 records (2 808 in 2020 vs 3763 in 2019) | Only a single comparison, no information on the usual variation of the number of records. |

### 9. Malaria

Here we present overview of the results mapping the indirect impact of COVID-19 on malaria as summarized in the main text. **S1 Table 14** shows an overview of all included studies with respect to reported outcomes, considered population and geographical area. Further tables compare projected and observed malaria cases (**S1 Table 15**), projected and observed malaria deaths (**S1 Table 16**) and the modelled impact of interruption of different malaria prevention measures and the observed interruptions (**S1 table 17**).

**S1 Table 14** : **Overview of studies related to malaria with respect to various characteristics.** Note that one study[37] reported both observational and modelled data. Shown are also general trends observed within the studied country/region: increase (↑), decrease (↓), changes in both directions (↕).

| | **Modelling studies (n=6+1)** | **Observational studies/reports/surveys (n=12+1)** |
|---|---|---|
| **Source** | Systematic search: 5[35, 37, 98-100], Grey literature: 1[101]<br>References: 1[39] | Systematic search: 10 [37, 102-110], Grey literature: 3 [54, 111, 112]<br>References: --- |
| **Region** | Africa: 6 [35, 37, 98-101] , 118 LIMCs: 1 [39] | Africa: 5 [37, 102, 104, 106, 110], Asia: 4 [105, 107-109], South America: 1 [103], Countries with malaria cases: 3 [54, 111, 112] |
| **Population** | General population: 6 [35, 37, 98-101], Pregnant women, children<5y: 1 [39] | General population: 11 [37, 54, 103-105, 107-112], Children < 5y: 2 [102, 106] |
| **Malaria cases** | 4: ↑ [37, 98, 100, 101] | 10: ↑ [107, 112] , ↕ [37, 106], ↓ [102, 103, 105, 108, 109, 111] |
| **Malaria deaths** | 6: ↑ [35, 37, 39, 98, 100, 101] | 3: ↕ [37], ↓ [109], --- [106] (too short observational time) |
| **Morbidity** | 3: ↑ [35, 37, 98] | --- |
| **Impact on malaria prevention measures** | 2: [39, 99] | 12 [54, 102-112] |
| **Adjustments of programs** | --- | 8 [54, 104-106, 108, 109, 111, 112] |

**S1 Table 15 Overview of projected and observed impact of COVID-19 on malaria cases.** ↑ = increase, ↓= reduction, ↕= both increase and reduction reported in the publication.

| Outcome | Modelling studies | Observational studies/reports/surveys | |
|---|---|---|---|
| | **Projected effect** | **Observed effect** | **Provided explanation** |
| **Malaria cases** | ↑ Africa:<br>• Uganda: + 877317 - 6561342[37]<br>• sub-Saharan Africa:<br>⇒ + 365860 - 4.4 million (+ 1 - 14 %) (countries without planned ITN campaigns in 2020) [101]<br>⇒ + 7.7 - 41.9 million (4 - 23 %) (countries with planned ITN campaigns in 2020) [101]<br>⇒ + 5 - 292 million (+ 2 - 113 %) [98]<br>⇒ + 9 - 46.4 million (+ 4 - 22 %) [100] | ↑ Asia:<br>⇒ Bhutan: + 7 (+ 17 %): increase in locally transmitted malaria cases from 2 to 20 (+ 36%) [107] | Pandemic-related interruption of prevention measures leads to an increase in locally transmitted malaria cases |
| | | ↑ Africa:<br>⇒ Botswana, Burkina Faso, Guinea, Madagascar, Malawi, Namibia, Mozambique, Sao Tome und Príncipe, Zambia, Zanzibar (Tanzania) [112] | No indication of dates or direct causalities |
| | | ↕ Africa:<br>• Uganda: + 374107 (+ 45 %, January 2020), + 344031 (+ 52 %, February 2020), - 44194 (- 6 %, March 2020) [37]<br>• Sierra Leone (1 district, children < 5 years): + 5 (+ 10 %) bis + 45 (+ 150 %, compared to January, February, March, May 2018 /2019), - 3 (- 8 %) up to – 27 (- 45 %, compared to April 2018/2019) [106] | Seasonal monthly fluctuations, if applicable |
| | | ↓ Africa:<br>• Sierra Leone (1 district, children < 5 years): - 85 (- 40 % clinical diagnosis), - 30 (- 25 % confirmed cases) [102]<br>• Kenia: decrease of 50% [111]<br>↓ Asia:<br>• Cambodia (three provinces, graphical representation): continuous decrease in number of cases since 2019 with increase in testing capacity. In April 2020, abrupt drop in the number of tests carried out, with continuously falling malaria case numbers [105]<br>↓ South America:<br>• Peru (1 region): reduction in 2020 from February to March by 22%, from March to April by 44%, from April to May by 88% and from May to 21 June by 99% [103] | Consequence of the COVID 19 pandemic |
| | | ↓ Asia:<br>• Sri Lanka: - 11 (- 32 %) [108]<br>• Myanmar: - 3923 (- 11 %) [109] | Good malaria prevention work |
| | | ↓ Africa:<br>• Angola, Benin, Burundi, Central African Republic, Chad, Comoros, Congo, Democratic republic of the Congo, Swaziland, Ghana, Nigeria, South Africa, Tanzania, Zimbabwe [112] | No indication of dates or direct causalities |

**S1 Table 16 Overview of projected and observed impact of COVID-19 on malaria deaths**. DALY = disability adjusted life-years lost, YLL = years of life lost, ITN = insecticide-treated bednets. ↑ = increase, ↓= reduction, →=no change, ↕= both increase and reduction reported in the publication.

| **Outcome** | **Modelling studies** | **Observational studies/reports/surveys** | |
|---|---|---|---|
| | **Projected effect** | **Observed effect** | **Provided explanation** |
| **Malaria deaths** | ↑ Africa:<br>• Uganda: + 3209 to 31046 [37]<br>• sub-Saharan Africa:<br>⇒ + 667 - 50515 (1 - 82 %) (countries without planned ITN campaigns in 2020) [101]<br>⇒ 24573 - 331630 (+ 8 - 102 %) (countries with planned ITN campaigns in 2020) [101]<br>⇒ + 25000 - 696000 (+ 5,9 -165 %) [98]<br>⇒ + 23590 - 382150 (6 - 99 %) [100]<br>⇒ East/ West Africa: + 79 - 2149 per million inhabitants [35] | ↓ Asia:<br>• Myanmar: - 3 (-27%) [109] | Good malaria prevention work |
| | | ↕ Africa<br>• Uganda: + 379 (+ 123 %, January 2020), + 455 (+ 243 %, February 2020), - 45 (- 19 %, March 2020) [37] | Seasonal monthly fluctuations, if applicable |
| | ↑ LMIC:<br>• + 28 - 152 (+ 1-2 %, maternal deaths, 118 countries) [39]<br>• + 90 - 1170 (+ 0 - 1 %, children under 5 years, 118 countries) [39] | → Africa:<br>• Sierra Leone (1 district, children < 5 years): 2 deaths until 5/2020 [106] | No statement possible yet, observation period too short |
| **Malaria associated morbidity (DALY, YLL)** | ↑ DALY: Uganda: + 257780 - 2427769 [37]<br><br>↑ YLL:<br>• East/ West Africa: + 4082 - 111097 per million inhabitants [35]<br>• sub-Saharan Africa: + 1 - 34 million [98] | / | / |

**S1 table 17 Projected consequences of interruption of malaria prevention measures on malaria cases and deaths and overview of observed disruptions of the measures.**

| Outcome | Modelling studies | Observational studies/reports/surveys |
|---|---|---|
| **Prevention measures** | • Interruption of mosquito net distribution campaigns causes the largest increase in malaria case numbers and malaria deaths (considered as a single measure) [35, 98].<br>• Reduction of access to effective malaria therapy causes the strongest increase in the number of malaria cases and the number of malaria deaths (considered as a single measure) [100, 101].<br>• The more prevention measures are reduced or interrupted at the same time, the greater the increase in the number of malaria cases [35, 98, 100, 101].<br>• The more prevention measures are simultaneously reduced or interrupted in the modelling, the greater the increase in malaria deaths [35, 39, 98, 100, 101].<br>• Modelling COVID-19 pandemic control scenarios: if restrictions are implemented for 12 months, there is the highest increase in malaria deaths if health systems are poorly managed [35, 98].<br>• The more the COVID-19 pandemic can be contained, the more mosquito nets can be distributed, the lower the malaria transmission [99].<br>• Reduction in health services could lead to a reduction in intermittent malaria prophylaxis in pregnancy [39]. | • Interruptions / reduction of prevention measures due to the COVID 19 pandemic or restrictions taken to combat the pandemic.<br>⇒ Logistic problems [54, 103, 104, 107, 110-112]<br>⇒ Supply bottlenecks [110-112]<br>⇒ Reduced headcount due to COVID-19 illness or pandemic deployment [54, 104, 107, 111, 112]<br>• Reduced use of health services [102, 103, 105, 110, 111]<br>• Reduced supply of or access to health services or hospitals [54, 103, 110-112]<br>• No pandemic-related impairment of prevention measures [106, 108, 109]<br>• Adaptation of prevention measures<br>⇒ Hygiene measures, provision of protective equipment [54, 104, 105, 109, 111, 112]<br>⇒ Staff training courses [104-106, 109, 111, 112]<br>⇒ Adaptation of the distribution concepts of the campaigns [54, 105, 111, 112]<br>⇒ Information campaigns [105, 109, 111, 112] |

### 10. HIV/AIDS

In this section we present additional figures and findings related to the indirect impact of COVID-19 on HIV prevention and treatment. **S1 Fig. 6** details reported reduction in HIV testing volumes and changes in positivity rates, all numerical values used in the plot can be found in **S1 Table 21**. **S1 Fig. 7** reports on changes in pre-exposure prophylaxis (PrEP) and **S1 Fig. 8** in post-exposure prophylaxis (PEP). Further, **S1 Table 19** collects the self-reported disruption in HIV services. Finally, to provide detailed numbers to the endpoints discussed in the paper, **S1 Table 20** details all the findings related to ART initiations, ART retention and engagement in care and **S1 Table 22** collects results of the modelling studies. In addition, assuming 20% loss to follow-up for 6 months and a subsequent return to 1990 mortality levels, with equal mortality across age-groups, and a 55% decline in ART initiations led to 475 319 excess DALYs(disability adjusted life-years) lost for Uganda[37]. For comparison, 405 100 excess DALYs were predicted for 20% incidence of COVID-19 and age-related mortality based on that of China. Please note that the tables are organised according to outcomes, so the same reference may appear several times.

**S1 Table 18 Overview of studies related to HIV with respect to various characteristics.** Two studies from Japan[113, 114] evaluated the same data, so we have dropped reporting on [114]. ART=antiretroviral therapy, PrEP = preexposure prophylaxis, PEP = postexposure prophylaxis

| | **Health care records (national or regional/ single hospitals or programmes)** | **Surveys** | **Grey literature** |
|---|---|---|---|
| **ART retention and engagement in care** | Europe: 0/2 [17, 115]<br>Americas: 0/2 [116, 117]<br>Africa: 1/0 [118] | Americas: 5 [119-123]<br>Africa: 2 [124, 125]<br>Asia: 2 [126, 127]<br>global: 3 [128-130] | Africa: 1 [131]<br>Asia: 1[132]<br>Europe: 1[133]<br>global: 3 [134-137] |
| **ART initiations** | Asia: 1/0 [138], Africa: 1/1[37, 118] | | global: 1 [135] |
| **Facility based HIV testing** | Africa: 1/1 [139]/[140]<br>Asia: 2/0 [113, 138]<br>Europe: 0/1[141]<br>Americas: 0/1[142]<br>Australia: 0/1[143]<br>Middle East: 0/ 1[144] | Americas: 3 [122, 145, 146]<br>Europe: 2 [79, 82] | Africa: 1[75]<br>Americas: 3 [147, 148](on PrEP),[149]<br>global: 1 [135] |
| **New diagnoses** | Asia: 3/0[113, 138, 150], Africa:0/1[37]<br>Europe: 0/2[115, 141]<br>Australia: 0/1[143] | | Europe: 3 [133, 151, 152] |
| **PrEP** | Africa: 0/1 [153]<br>Australia: 0/1[154] | Americas: 4[122, 123, 145, 146]<br>Europe: 1[155]<br>Australia: 2 [156, 157] | Americas: 3[147-149] |
| **PEP** | Europe: 0/2[158, 159]<br>Australia: 0/1[160]<br>Middle East: 0/1[144] | | |
| **Disruption and uptake of services in general** | Asia: 1/0[113]<br>Africa: 1/0[161] | Asia: 1[162] ,<br>Americas: 1[163]<br>Europe: 1[164], global: 1[50] | Americas: 1[149]<br>Europe: 3[133, 151, 152]<br>global: 1[54, 165] |
| **Other prevention services** | Africa: 0/2[140, 166] | | Africa: 1[167] |
| **Modelling studies** | Incidence and/or mortality | Africa: 5 [35, 36, 42, 43] culminated in comparison in [168]<br>USA: 3 [38, 169, 170]<br>China: 1 [171] | |
| | Disability adjusted life years lost | Africa: 1 [37] | |

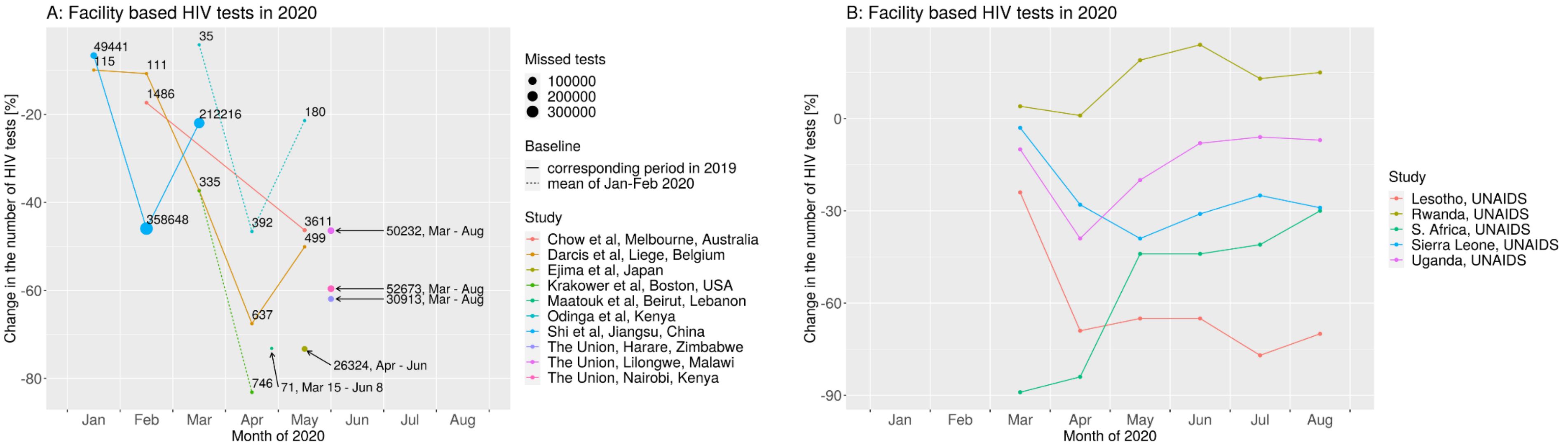

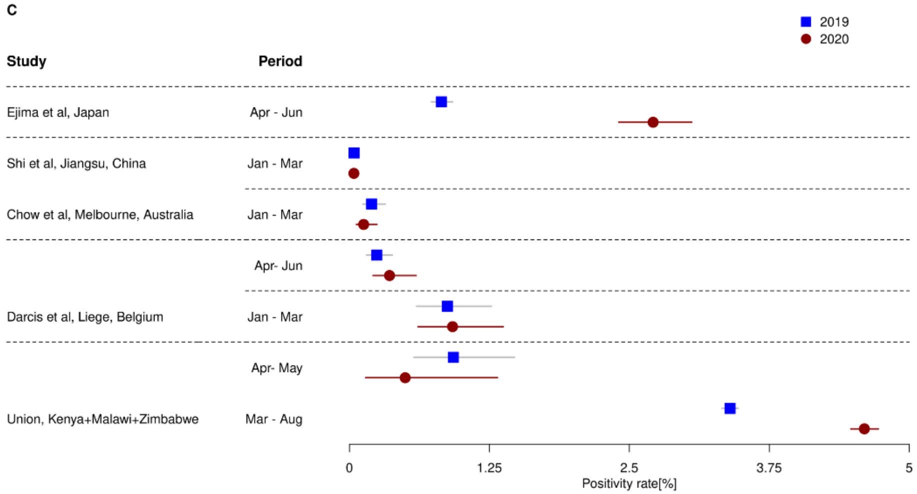

A. Changes in facility-based HIV testing volumes. Shown are the % and absolute decrease in the number of HIV tests. Note that using the mean of Jan-Feb 2020 as the baseline does not account for possible seasonality. The WHO declared COVID-19 to be a pandemic on March 11, 2020, whereas in China the lockdown measures were introduced already in late January 2020. The 85% reduction reported by Krakower et al in April 2020 relates to HIV testing on PrEP and can be compared to 59% of 189 PrEP prescribers in the USA reporting skipping HIV testing on PrEP refill [148].

B. Percent change in HIV testing volumes as read off the plots reported by UNAIDS[135]. In this case the comparison is made to the average of Jan-Feb 2020, so the results may be confounded by seasonality. For example, if we calculate the same percent change for March–August 2019 relative to the average of Jan-Feb 2019 from data reported for one centre in Liege, Belgium[141], we would obtain a decrease between 7.5 to 20% for the whole period.

C. Positivity rate as observed by the different studies in 2020 and 2019. Shown are the point estimates and 95% confidence intervals based on the Agresti-Coull method for binomial proportions.

**S1 Fig. 6 Reported reduction in HIV testing volumes and changes in the positivity rate.** The underlying numbers can be found in **S1 Table 21**.

PrEP

S1 Fig. **7** shows the self-reported decrease in usage of PrEP and difficulties accessing PrEP. The main reasons for stopping PrEP in Brazil were impediments with refill (47%), followed by sex abstinence (40%). In all other studies the main reason for stopping or reducing PrEP seemed to be a lower perceived HIV risk/decrease in sexual activities with casual partners. Problems accessing PrEP as a reason for stopping were reported only by 7%[156] , 8% [148], 11% [157]. A single centre in Boston [147] reported a decline in PrEP patients, PrEP initiations and a large increase in PrEP refill lapses compared to the mean of January-February 2020(3188 patients, 107 initiations, 141.5 lapses). Namely, in March 2020, there were 2% fewer patients, 44% fewer initiations and 44% more lapses, whereas in April 2020, there were 6% fewer patients, 68% fewer initiations and 187% more lapses. From Australia, a 24% decrease in the mean weekly number of prescriptions (Apr-June 2020 vs Jan 2019-March 2020) was reported[154] and a specific programme in South Africa observed a 68% increase in the rate of missed PrEP visits of pregnant women (from 34% of missed visits pre-lockdown to 57% of missed visits during lockdown) [153].

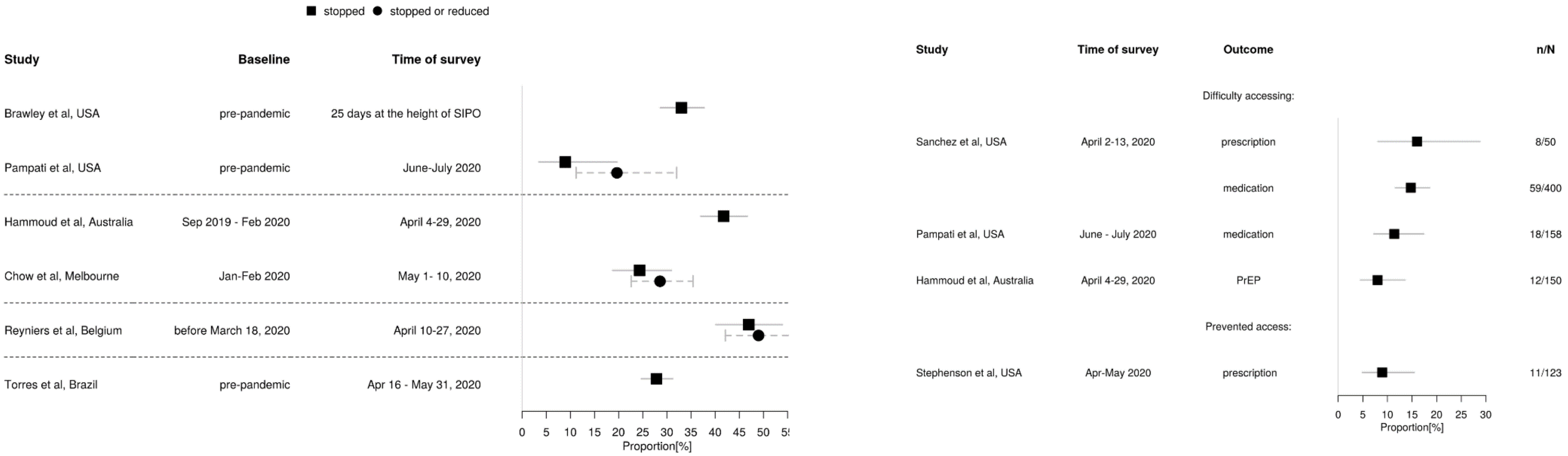

**S1 Fig. 7 Impact on PrEP usage.** Left: Proportions of respondents reporting changes in PrEP use. Shown are also the 95% confidence intervals based on the Agresti-Coull method. Right: Proportions of respondents reporting difficulties accessing PrEP. Shown are also the 95% confidence intervals based on the Agresti-Coull method. For comparison, 25% of 169 Ryan White grantees (USA) reported a decreased ability to provide PrEP, and 47% reported a decreased demand for it during the pandemic (until August 2020) [149], whereas in another survey[148] 95% of 189 U.S. prescribers declared the ability to prescribe PrEP also during the shelter-in-place order. For completeness, here are the absolute numbers underlying the left plot: Brawley et al: 135/409, Pampati et al: 5/56 and 11/56, Hammoud et al: 167/400, Chow et al: 46/189 and 54/189, Reyniers et al: 93/198 and 97/198, Torres et al: 204/733.

*PEP*

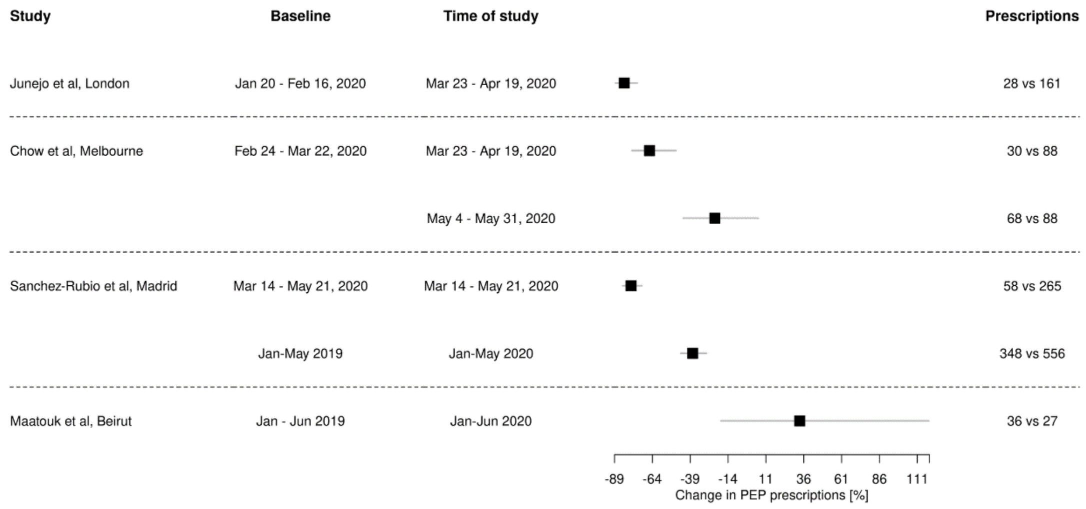

**S1 Fig. 8 Reduction in PEP prescriptions.** Shown are 95% confidence intervals based on the assumption of a Poisson distribution for the observed counts. Although the numbers were not reported, Sanchez-Rubio et al. [159] commented that the numbers started to increase even before the lockdown was at the end, which is what Chow et al. [160] observed as well.

*Other prevention services*

Compared to the mean outreach contacts of 5016 in 3 centres in Kenya [140] in Jan-Feb 2020, the programme registered a decrease of 2%(March 2020), 30%(April 2020) and 13%(May 2020) in the outreach contacts. Similarly, an assisted partner notification programme in the same country observed a decrease of 8%(March 2020) and of 36%(Apr 2020) in female index enrolments as compared to the monthly mean of 160 enrolments in Jan-Feb 2020, across two centres involved[166]. Regarding VMMC, countries reported decreased numbers of circumcisions in April, May, June[167] than in January 2020(most notably Lesotho, South Africa, Mozambique, Botswana and Zimbabwe). Without any quantification, another report of UNAIDS[135] mentions that by late 2020, the VMMC programmes in Kenya, Rwanda, Botswana and South Africa were returning to normal.

*Disruption and uptake of HIV services in general*

**S1 Table 19** shows the self-reported service interruptions. In addition, a decrease of overall outpatients visits was noted by several authors: a decrease of 64% in the second quarter of 2020 in Japan[113], of around 60% in both March-April 2020 and May-June 2020 in one centre in Greece[152], always compared to the corresponding period in 2019. In Croatia, the weekly mean went from 46(February 2020) to 22.5(March 2020)[133]. In contrast, an immediate increase in the mean visits/clinic /day of 8.0 (95%CI: 2.3-13.7) following the introduction of level 5 lockdown was observed in KwaZulu Natal[161]. This increase was followed by a slight steady decrease until the start of level 4 lockdown, when another stepwise increase of 11 visits/clinic/day (95% CI: 4.2 – 17.8) was observed. Without being specific about the qualitative assessment, as of June 1, 2020, The Global Fund reported 18% of 106 countries experiencing high to very high disruption to HIV services, and 66% experiencing moderate disruptions[54]. The situation seemed to improve only very slightly, with still 55% experiencing moderate disruptions in December [165].

**S1 Table 19 Self-reported HIV service interruptions.**

| Country | Disruption | Country | |
|---|---|---|---|
| Guangxi, China | Of 1029 professionals [162]:<br>• 59.5% report shortage in personnel<br>• 48.7% report conflict between Covid and HIV tasks<br>• 25% report their clinic has no capacity to maintain HIV services | Europe | Of 19 countries represented by single individuals in March 2020 [164]:<br>• 16%: shorter hours or reduced staff<br>• 53%: only ART distribution(no visits, no blood tests)<br>• 58%: physicians sharing Covid-19 duties<br>Of 9 HIV clinics in Greece[151] asked in June 2020:<br>• 25% cancelled scheduled visits during lockdown completely<br>• 50% covered only emergencies and prescriptions during lockdown |
| Ryan White grantees, USA | Of 27 clinics in South Carolina in March 2020[163]:<br>• 26% : complete interruption<br>• 56% partial interruption<br>Of 169 grantees in the USA covering the period up to Sep 2020[149]:<br>• 28%: shutting down most or all HIV prevention services<br>• 5%: shutting down most or all HIV treatment services | Global | Of 346 professionals[50] interviewed in May-August 2020:<br>• 16%: experienced serious* challenges in provision of standard treatment/medicine<br>• 33% experienced light** challenges in provision of standard treatment/medicine<br>• 24%: experienced serious* challenges in provision of routine diagnostic services<br>• 36%: experienced light* challenges in provision of routine diagnostic services<br>* it is much harder, it is very difficult or impossible<br>**it is slightly harder |

**S1 Table 20 Overview of studies reporting on ART retention and engagement in care.** PLWH = people living with HIV, SUD = substance use disorder, MSM = men who have sex with men, AMIS = American Men's Internet Survey, RCT = randomized controlled trial, TGNB = transgender, non-binary individuals

| Detailed outcome | Country | Population | Period | Baseline | Result | Author |
|---|---|---|---|---|---|---|
| **ART adherence** | | | | | | |
| Electronically measured adherence | USA (Los Angeles, California) | Black population living with HIV (from a parent RCT studying motivational interviewing intervention to improve ART adherence) | May -July 2020 | Previous 1-month measurement closest to the COVID-19 survey for each participant | OR 0.59 for good adherence (i.e. >80% doses taken per 1 additional experienced negative impact of the pandemic) | Bogart et al[119] |
| 3-item self-reported instrument for retrospective adherence converted into scale 0-100 | USA (Atlanta, Georgia) | PLWH 20-37 years old screened positive for substance use (participants of ART adherence study) | Month after onset of COVID-19 protective actions (median 9 months in the study) | Previous month | Significant improvement in the mean adherence score in the month after onset of COVID-19 protective actions (from 52.8 to 77.6) | Kalichman et al[120] |
| Missed ART dose on 2 or more days of the week | USA (Wisconsin) | PLWH, SUD at high risk of treatment failure | Mar 24 – May 4, 2020 | Jan 31 – Mar 12, 2020 | OR 2.81 during pandemic, p=0.084 | Hochstatter et al[121] |

| | | | | | | |
|---|---|---|---|---|---|---|
| Difficulties with ART refill (possibly including inability to refill) | USA | MSM (AMIS participants) | Apr 2 – 13, 2020 | --- | 8/121 =7% | Sanchez et al[122] |
| | Global (Belgium 32.2%, Brazil 28.1%, Eastern Europe 18.6%) | PLWH (online questionnaire) | Apr 9 – May 17, 2020 | --- | 29/317=9% | Siewe Fodjo et al[129] |
| | Global(mostly Brazil, France, Mexico, Taiwan, Russia) | Cisgender gay men and other MSM (networking app Hornet) | Apr 16 – May 4, 2020 | --- | 84/455=18% | Santos et al[128] |
| | Brazil | MSM and TGNB (Hornet, What's App, facebook) | Apr 16 – May 31, 2020 | --- | 138/800=17% | Torres et al[123] |
| Limited access to ART | 138 countries (Russia, Turkey, France, Brazil,…) | LGBT+(online) | Apr 15 – May 18, 2020 | --- | 21% (from around 2200-2300) | Lamontagne et al[137] |
| Unable to get ART refill | Western Kenya | Adolescent (10-24 years) living with HIV (ongoing cohort study to improve outcomes) | 10 first weeks of COVID-19 | --- | 15/486=3% | Dyer et al[124] |
| | Global(mostly Brazil, France, Mexico, Taiwan, Russia) | Cisgender gay men and other MSM (networking app Hornet) | Apr 16 – May 4, 2020 | --- | 20/455=4% | Santos et al[128] |
| | USA (Los Angeles, California) | Black population living with HIV (from a parent RCT studying motivational interviewing intervention to improve ART adherence) | May – July 2020 | --- | 6/101=6% | Bogart et al[119] |
| No access to ART | 138 countries (Russia, Turkey, France, Brazil,…) | LGBT+(online) | Apr 15 – May 18, 2020 | --- | 7% (from around 2200-2300) | Lamontagne et al[137] |
| Decreased ability to adhere to ART | USA | MSM (AMIS participants) | Apr 2 – 13, 2020 | Pre-covid | 6/121=5% | Sanchez et al[122] |
| | Uganda | HIV positive adults (RCT participants in Mildmay Uganda clinic) | Apr 6 – 17, 2020(personal communication) | Pre-covid | 14/100=14% | Linnemayr et al[125] |
| | Global (Belgium 32.2%, Brazil 28.1%, Eastern Europe 18.6%) | PLWH (online questionnaire) | Apr 9- May 17, 2020 | Pre-covid | 11/312=4% | Siewe Fodjo et al[129] |
| Risk of ART interruption = having not enough medication under current traffic and movement restrictions | China | PLWH on ART (online questionnaire) | Feb 5-10, 2020 | ---- | 331/1014=33% | Guo et al[127] |

| | | | | | | |
|---|---|---|---|---|---|---|
| Risk of ART interruption = medication for less than 10 days without a clear way for refill | China | PLWH (recruited through HIV-themed WeChat public accounts) | Feb 5-17, 2020 | ---- | 917/5084=18% | Sun et al[126] |
| Disruptions in the number of people on ART | Global | PLWH on ART (country programme data reported to HIV Services Tracking Tool) | Mar – Sep 2020 | Jan – Feb 2020 | 16/25 countries no disruptions, some have seen drops in the number of people on ART: Peru, -14%, July 2020 South Africa: -5/-6% in July/August 2020, Dominican Republic: up to -10% Apr – Sep 2020 | UNAIDS[135] |
| **Viral load suppression** | | | | | | |
| Unsuppressed viral load | USA (San Francisco) | PLWH attending a safety net clinic (Ward 86) | Apr 1 – Apr 30, 2020 | Dec 1, 2019 – Feb 29, 2020 | OR 1.31 (1.08– 1.53), n=1766 | Spinelli et al[116] |
| | USA (San Francisco) | Homeless PLWH in a low-barrier HIV primary care programme (Ward 86) | Mar 17 – Aug 16, 2020 | Oct 17, 2020 – Mar 16, 2020 | OR 1.19 (0.78-1.82), n=66 | Hickey et al[117] |
| Number of avireamic patients | Italy (Rome) | HIV patients (HIV/AIDS unit of S. Gallicano Dermatological Institute) | Mar 9 – Aug 20, 2020 | Pre-lockdown | 95.8% of 506 aviraemic before lockdown remained avireamic | Giuliani et al[17] |
| **ART dispensation** | | | | | | |
| Pick-up coverage | Lagos, Nigeria | Clients of Centre for Integrated Health Programme | Apr – May 2020 | --- | 97% of all eligible for pick-up received ART refill at the end of the lockdown period thanks to adaptation of services | Ogirima et al[131] |
| Number of ART dispensations | Italy (Brescia) | HIV patients (tertiary referral center for Infectious Diseases and HIV in Brescia) | Mar-Apr 2020 | Oct – Nov 2019 | -23.1% (-33.6% in March, -12.9% in April) | Quiros-Roldan et al[115] |
| Weekly number of individuals receiving treatment | Nepal | Patients in sites supported by LINKAGE project and other partners | Mar 22 – May 16, 2020 | Jan 26 – Mar 21, 2020 | Ca 800-1200 vs ca 1700-2000(baseline) | UNAIDS[134] |
| **ART consultations** | | | | | | |
| Number of ART consultations | Myanmar | ART patients in a primary care clinic | Apr 2020 | Apr 2019 | 6630 vs 6009 (baseline) | Tun et al[132] |
| **Viral load tests** | | | | | | |
| Number of viral load tests | Croatia | PLWH treated in Croatia | Mar- Jun 2020 | Mar –Jun 2019/2018 | ca -30% (545 vs 777/762) | Bogdanic et al[133] |

| | | | | | | |
|---|---|---|---|---|---|---|
| | South Africa | Tests carried out by the sole laboratory service provider to public sector | Mar 27 – Apr 30, 2020/May 1 – June 1, 2020 | Feb 1 – Mar 26, 2020 | -22%/-5% in weekly averages | Madhi et al[118] |
| **ART initiations** | | | | | | |
| Number of ART initiations | China(Jiangsu) | PLWH starting ART (data from Comprehensive response Information Management System for HIV/AIDS in China) | 1st quarter 2020 | 1st quarter 2019 | -34% (208 vs 315) | Shi et al[138] |
| More sustained disruptions in ART numbers | Global | PLWH starting ART (country programme data reported to HIV Services Tracking Tool) | Mar – Aug 2020 | Jan-Feb 2020 | 21/28 countries, individual monthly changes in the range: -45%(Sierra Leone) - +88% (Nigeria) | UNAIDS[135] |
| Number of ART initiations(weekly mean) | Uganda | PLWH starting ART (recorded by PEPFAR, Uganda ministry of Health and partners) | Apr 2020 (weeks 14-16) | Oct 2019 – Mar 2020 | -60% ( 1096 vs 2721) [reported also -37% in week 23, June 2020] | Bell et al[37] |
| CD4+ tests (proxy for initiation) | South Africa | Tests carried out by the sole laboratory service provider to public sector | Mar 27 – Apr 30, 2020/May 1 – June 1, 2020 | Feb 1 – Mar 26, 2020 | -33%/ -15% in weekly averages | Madhi et al[118] |
| **Keeping HIV appointments** | | | | | | |
| Rate of missed visits | USA (San Francisco) | PLWH attending a safety net clinic (Ward 86) | Apr 1 – 30, 2020 | Dec 1, 2019 – Feb 29, 2020 | 599/1997(30%) vs 1287/4153(31%, baseline), OR 0.91 (0.77-1.09) | Spinelli et al[116] |
| | Italy (Brescia) | HIV patients (tertiary referral center for Infectious Diseases and HIV in Brescia) | Mar-Apr 2020 | Oct – Nov 2019 | 94/1162(8%) vs 60/1217(5%, baseline) | Quiros-Roldan et al[115] |
| Keeping regular visits (visit each month) | USA (San Francisco) | Homeless PLWH in a low-barrier HIV primary care programme (Ward 86) | Mar 17 – Aug 16, 2020 | Oct 17, 2020 – Mar 16, 2020 | OR 0.66 (0.39-1.11) | Hickey et al[117] |
| Confidence to keep the next HIV appointment (scale 1-7) | USA (Wisconsin) | PLWH, SUD at high risk of treatment failure | Jan 31 – Mar 12, 2020 | Mar 24 – May 4, 2020 | Decrease in mean score from 6.89 to 6.5, p=0.021(Poisson regression) | Hochstatter et al[121] |
| **ART services interruptions** | | | | | | |
| Disruption to services | Global (all Regions, but the Region of the Americas) | Data collected by WHO regional offices through facilitated discussion with key informants within the national ministry of health | May-Jul 2020 | Pre-covid, unspecified | partial disruption(5-50% pts not treated as usual): 31% of 101<br>sever disruption(>50% of pts not treated as usual): 1% of 101 | WHO[136] |

| | | | | | | |
|---|---|---|---|---|---|---|
| **Difficulties accessing HIV provider** | | | | | | |
| Unable to access provider | Western Kenya | Adolescent (10-24 years) living with HIV (ongoing cohort study to improve outcomes) | First 10 weeks of COVID-19 | --- | 85/486 =17% | Dyer et al[124] |
| | Global(mostly Brazil, France, Mexico, Taiwan, Russia) | Cisgender gay men and other MSM (networking app Hornet) | Apr 16 – May 4, 2020 | --- | 111/473 =23% | Santos et al[128] |
| | Global (about one third from Russian Federation) | PLWH (users of Hornet, gay social networking app) | Apr 16 – May 24, 2020 | --- | 218/1105=20% | Rao et al[130] |
| Trouble making/keeping appointment | USA | MSM (AMIS participants) | Apr 2 – 13, 2020 | --- | 24/121=20% | Sanchez et al[122] |
| COVID-19 impacts my ability to come to clinic | Uganda | HIV positive adults (RCT participants in Mildmay Uganda clinic) | Apr 6 – 17, 2020 | --- | 76/100=76% | Linnemayr et al[125] |

**S1 Table 21 Overview of findings related to HIV testing and new diagnoses.** MSM=men who have sex with men

| **Detailed outcome** | **Country** | **Population** | **Period** | **Baseline** | **Result** | **Author** |
|---|---|---|---|---|---|---|
| **Facility based HIV tests** | | | | | | |
| Change in the number of tests | Australia (Melbourne) | Data from Melbourne Sexual Health Centre | 1st quarter \| 2nd quarter of 2020 | 1st quarter \| 2nd quarter of 2019 | -17% (7083 vs 8569) \| -46% (4187 vs 7798) [comparison between the respective quarter of 2020 and 2019] | Chow et al[143] |
| | Belgium (Liege) | Data from Liege University Hospital | Jan \| Feb \| Mar \| Apr\| May 2020 | Jan \| Feb \| Mar \| Apr\| May 2019 | -10% (1046 vs 1161) \| -11%(925 vs 1036) \| -37% (634 vs 1011) \|-68% (306 vs 943)\| -50% (497 vs 996)<br>[comparison between the respective month in 2020 and 2019] | Darcis et al[141] |
| | Lebanon (Beirut) | Patients at Clemenceau Medical Center | Mar 15 - Jun 8, 2020 | Mar 15 - Jun 8, 2019 | -73% (26 vs 97) | Maatouk et al[144] |
| | Kenya (Kisumu, Kiambu, Momabasa) | combined data from 3 prevention programmes aimed at MSM | Mar \| Apr \| May 2020 | Jan-Feb 2020 | -4% (806 vs 841) \| -47%(449 vs 841) \| -21% (661 vs 841) [comparison between the respective month in 2020 and monthly mean of Jan-Feb 2020] | Odinga et al[140] |

| | | | | | | |
|---|---|---|---|---|---|---|
| | Kenya | Data from 18 health facilities in Nairobi | Mar-Aug 2020 | Mar-Aug 2019 | -60% (35676 vs 88349) | Union[75] |
| | | Data from 8 health facilities in Lilongwe | Mar-Aug 2020 | Mar-Aug 2019 | -46% (57972 vs 108204) | Union[75] |
| | Zimbabwe | Data from 10 health facilities in Harare | Mar-Aug 2020 | Mar-Aug 2019 | -62% (18987 vs 49900) | Union[75] |
| | China(Jiangsu) | Screening tests, data from Comprehensive response Information Management System for HIV/AIDS in China | Jan \| Feb \| Mar 2020 | Jan \| Feb \| Mar 2019 | -7% (699310 vs 748751) \| -46% (422040 vs 780688)\| -22% (754335 vs 966551) [comparison between the respective month in 2020 and 2019] | Shi et al[138] |
| | Japan | Data from quarterly reports from the National AIDS surveillance Committee | 2nd quarter of 2020 | 2nd quarter of 2019 | -73% (9584 vs 35908) | Ejima et al[113] |
| | Lesotho | country programme data reported to HIV Services Tracking Tool | Mar \| Apr \|May\| Jun\| Jul \| Aug 2020 | Jan-Feb 2020 | -24% \| -69% \| -65% \| -65% \| -77% \| -70% [comparison between the respective month in 2020 and monthly mean of Jan-Feb 2020] | UNAIDS [135] (approximately read off the plots) |
| | Sierra Leone | | | | -3% \| -28%\| -39%\| -31%\| -25%\| -29% [comparison between the respective month in 2020 and monthly mean of Jan-Feb 2020] | |
| | South Africa | | | | -89%\|-84%\|-44%\|-44%\|-41%\|-30% [comparison between the respective month in 2020 and monthly mean of Jan-Feb 2020] | |
| | Uganda | | | | -10%\|-39%\|-20%\|-8%\|-6%\|-7% [comparison between the respective month in 2020 and monthly mean of Jan-Feb 2020] | |
| | Rwanda | | | | 4%\|1%\|19%\|24%\|13%\|15% [comparison between the respective | |

| | | | | | | |
|---|---|---|---|---|---|---|
| | | | | | month in 2020 and monthly mean of Jan-Feb 2020] | |
| Change in the number of HIV tests on PrEP | USA(Boston, MA) | Data from Ben Israel Deaconess Medical Center | Mar \|Apr 2020 | Jan – Feb 2020 | -37% (562 vs 897) \| -83% (151 vs 897) [comparison of the respective month in 2020 with monthly mean of Jan-Feb 2020] | Krakower et al[147] |
| Uptake of HIV testing at birth | South Africa (KwaZulu-Natal) | Data from District Health Information System | Mar \| Apr \| May \| Jun 2020 | Jan –Feb 2020 | Uptake of 89.6%\| 88.2%\| 80.1%\|91.1%, baseline: monthly mean 93.9% 95% CI: 92.4 – 95.4 | Jensen et al[139] |
| HIV testing rate in emergency department | USA (Chicago,Illinois) | Data from Emergency Dept., University of Chicago | Mar 5 – Apr 18, 2020 | Jan 1 – Mar 4, 2020 | 1789/8616(21%) vs 3247/14524(22%, baseline), no significant difference | Stanford et al[142] |
| Being prevented from HIV testing | USA | Gay, bisexual and other MSM(recruited through facebook, Instagram, Grindr app) | Apr – May 2020 | Pre-pandemic | 32% of 518 | Stephenson et al [145] |
| Experienced difficulties getting tested when tried | USA | MSM (AMIS participants) | Apr 2 – 13, 2020 | --- | 18% of 286 | Sanchez et al[122] |
| | USA | MSM on PrEP(recruited online for a cohort study examining trajectories of PrEP use, Oct 2019 – Jul 2020) | Jun-Jul 2020 | --- | 24% of 45 | Pampati et al[146] |
| Decreased demand for testing (since the beginning of pandemic) | USA | Ryan White HIV/AIDS Program medical provider grantees | Aug 2020 | --- | 64% of 169 | KFF[149] |
| Decreased ability to test (since the beginning of pandemic) | | | | | 61% of 169 | |
| Skipping HIV testing on PrEP refill | USA | PrEP providers from American Academyof HIV Medicine’s database | 25-day period of shelter-in-place orders implementation | --- | 59% of 189 providers | Brawley et al[148] |
| | WHO European region | | Mar-May 2020 | Mar - May 2019 | 60% of 86 | Simoes et al[79] |

| | | | | | | |
|---|---|---|---|---|---|---|
| Severe reduction(>50%) in HIV testing volumes | | Laboratories, primary and secondary level care clinics, community sites, national level public health institutions, ministries of health (contacted through EuroTEST consortium network) | Jun-Aug 2020 | Mar – May 2019 | 20% of 86 | |
| HIV testing rates per 1000 service users | Spain (Basque Country, Catalonia, Madrid, Valencia) | Harm reduction centres | Mar – Jun 2020 | Mar – Jun 2019 | Very heterogeneous, in Apr 2020 drop to 0 for 4 of 5 centres (from 9.5 -211.9 in Apr 2019), in May and June 2020 both increase and decrease as compared to May and June 2019 | Picchio et al[82] |
| **New diagnoses** | | | | | | |
| Change in the number of new cases | China(Jiangsu) | data from Comprehensive response Information Management System for HIV/AIDS in China | 1st quarter of 2020 | 1st quarter of 2019 | -258 cases (-24%) | Shi et al[138] |
| | Japan | Data from quarterly reports from the National AIDS surveillance Committee | 2nd quarter of 2020 | 2nd quarter of 2019 | -35 cases (-12%) | Ejima et al[113] |
| | Taiwan | Data from Taiwan's Centre for Disease Control | Jan-Aug 2020 | Jan-Aug 2019 | -293 cases (-23%) | Chia et al[150] |
| Monthly median of new cases | Belgium (Liege) | Data from Liege University Hospital | Apr – May 2020 | Jan 2019 – Mar 2020 | 2 vs 8 (baseline) | Darcis et al[141] |
| Monthly mean number of new cases | Italy (Brescia) | HIV patients (tertiary referral center for Infectious Diseases and HIV in Brescia) | Mar-Apr 2020 | 2019 | 2.5 vs 6.4 (baseline) | Quiros-Roldan et al[115] |
| Weekly mean of positive tests | Uganda | Data from PEPFAR, Uganda ministry of Health and partners | Apr 2020 (weeks 14-16) | Oct 2019-Mar 2020 (up to week 13) | 1284 vs 3224(baseline) = -60% [reported is -37% decline in June 2020, week 23] | Bell et al[37] |
| Number of new cases | Australia(Melbourne) | | 1st quarter of 2020 | 1st quarter of 2019 | 9 vs 17(baseline) | Chow et al[143] |

| | | | | | | |
|---|---|---|---|---|---|---|
| | | Data from Melbourne Sexual Health Centre | 2nd quarter of 2020 | 2nd quarter of 2019 | 15 vs 19(baseline) | |
| | Croatia | Data from University Hospital for Infectious Diseases, Zagreb, treating all PLWH in Croatia | Mar – Jun 2020 | Mar –Jun 2019 and 2018 | 12 vs 22(in 2019) or 28(in 2018) | Bogdanic et al[133] |
| | Greece | Information from an expert panel of 9 HIV clinicians | Mar-May 2020 | Jan-Feb 2020 | Approx. 50% drop | Lourida et al[151] |
| | Greece(Alexandroupolis) | Data from HIV Clinic of Alexandroupolis | 1st semester 2020 | 1st semester 2019 | 13 vs 6 (baseline) (diagnoses in 2020 mainly after lockdown) | Panagopoulos et al[152] |
| **HIV positivity rates(number of new HIV diagnoses as a percentage of the number of HIV tests done)** | | | | | | |
| | Japan | Data from quarterly reports from the National AIDS surveillance Committee | 2nd quarter of 2020 | 2nd quarter of 2019 | 260/9584(2.7%) vs 295/35908(0.8%) | Ejima et al[113]* |
| | China(Jiangsu) | data from Comprehensive response Information Management System for HIV/AIDS in China | 1st quarter of 2020 | 1st quarter of 2019 | 749/1875685 (0.04%) vs 1057/2495990 (0.04%) | Shi et al[138] |
| | Australia (Melbourne) | Data from Melbourne Sexual Health Centre | 1st quarter of 2020 | 1st quarter of 2019 | 9/7083 (0.13%) vs 17/8569 (0.2%) | Chow et al[143] |
| | | | 2nd quarter of 2020 | 2nd quarter of 2019 | 15/4187 (0.36%) vs 19/7798(0.24%) | |
| | Belgium (Liege) | Data from Liege University Hospital | Jan –Mar 2020 | Jan-Mar 2019 | 24/2605(0.92%) vs 28/3198 (0.88%) | Darcis et al[141] |
| | | | Apr-May 2020 | Apr-May 2019 | 4/803(0.50%) vs 18/1939(0.93%) | |
| | Kenya, Zimbabwe, Malawi | 18 centres in Nairobi, 9 in Harare, 8 in Lilongwe | Mar-Aug 2020 | Mar-Aug 2020 | 5181/112635 (4.6%) vs 8379/246453 (3.4%) | Union[75]** |

*Calculated from the reported numbers on HIV case with and without AIDS diagnoses. **The absolute numbers are calculated from the percentages.

**S1 Table 22 Overview of the impact of individual HIV service/care interruptions on HIV-incidence and mortality as reported by the modeling studies.** From studies related to African countries[35, 36, 42, 43, 168], we report only on the common comparison in [168]. For comparison, one study [38] considered 50% disruption in services(ART initiation, ART engagement, HIV testing, PrEP uptake, syringe service programme, medication for opioid users) over 12 months starting in March 2020 only combined and observed increase in HIV incidence over 2020-2025 in the range 7-16% for six US cities.

| | % increase in HIV-incidence | | | | % increase in HIV-related mortality | | |
|---|---|---|---|---|---|---|---|
| | 1-year incidence | | | 5-year incidence | 1-year mortality | | |
| | Jewell[168], 50% disruption over 6 months, sub-Saharan Africa | Booton[171], 50% disruption over 3 months, 4 cities in China | Mitchell[169], 50% disruption over 6 months, MSM in Baltimore | Jenness[170], 50% disruption for 18 months, MSM in Atlanta area | Jewell[168] , 50% disruption over 6 months, sub-Saharan Africa | Booton[171], 50% disruption over 3 months, 4 cities in China, | Mitchell[169], 50% disruption over 6 months MSM in Baltimore |
| ART interruption | 1-16%, 126% | | | | 39-87% | | |
| ART retention | | | | 9% | | | |
| Condom use | 3-19% | 19% | 10% | | 0% | 0.5% | 0% |
| PMTCT | 2-5% | | | | 1-3% | | |
| HIV testing | 1% | 1.8% | 1% | | 0-1% | 0.1% | 0.1% |
| HIV screening | | | | 3.2% | | | |
| Viral load testing | 0-3% | | | | 0-4% | | |
| ART initiation | 0-2% | 2.4% | 1.1% | | 0-3% | 2.5% | 5% |
| VMMC | 0% | | | | 0% | | |
| Increase in death rate due to overwhelmed health care | 0% | | | | 2-6% | | |
| Viral suppression | | 15% | 29% | | | 18% | 46% |
| PrEP initiations | | | | 2.6% | | | |
